# A rapid evidence map of what evidence is available on the effectiveness of community diagnostic centres

**DOI:** 10.1101/2022.12.01.22282959

**Authors:** Alesha Wale, Chukwudi Okolie, Jordan Everitt, Amy Hookway, Hannah Shaw, Kirsty Little, Ruth Lewis, Alison Cooper, Adrian Edwards

## Abstract

The COVID-19 pandemic has resulted in increased demand and delays to diagnostic services. Community diagnostic centres (which are generally referred to as Regional Diagnostic Hubs in Wales) aim to reduce this backlog and the waiting times for patients by providing a broad range of elective diagnostic services in the community, away from acute hospital facilities. As diagnostic services account for over 85% of clinical pathways and cost the National Health Service (NHS) over six billion pounds a year (NHS 2022), community diagnostic centres across a broader range of diagnostic services may be an effective, efficient, and cost-effective introduction to the UK health sector. This Rapid Evidence Map aimed to identify, describe, and map the available evidence on the effectiveness of diagnostic centres. 50 primary studies were identified. Studies were published between 1995 and 2021: A wide range of study designs were used, and studies were conducted in a range of countries including the UK. 30 studies were specific to cancer diagnosis, whilst the remaining 20 studies focused on diagnosis associated with: anaemia, autism, cerebral palsy, intellectual disability, multiple sclerosis, respiratory conditions, shoulder pain, and unexplained fever Eleven studies reported information on multi-condition diagnostic centres, rather than a specific condition.

The majority of studies were conducted within hospital settings. Two studies evaluated diagnostic centres within a community setting. The diagnostic centres offered a wide range of diagnostic tests and incorporated different staff and facilities. Participants were mainly referred by GPs, primary care centres and emergency departments. However, referrals were also made from outpatient clinics located within the same hospital as the diagnostic centre.

Over 100 different outcomes were reported covering: patient data and referral outcomes, clinical outcomes, performance outcomes, economic outcomes, and patient and physician-reported outcomes.

The findings of this rapid evidence map were used to select a substantive focus for a subsequent rapid review on community diagnostic centres that can be accessed by primary care teams.

### Rapid Review Details

#### Review conducted by

Public Health Wales (PHW)

#### Review Team

- Alesha Wale, Public Health Wales,
- Chukwudi Okolie, Public Health Wales,
- Jordan Everitt, Public Health Wales,
- Amy Hookway, Public Health Wales,
- Hannah Shaw, Public Health Wales,
- Kirsty Little, Public Health Wales

**Review submitted to the WCEC on:** October 2022

**Stakeholder consultation meeting:** 13^th^ September 2022

**Rapid Evidence Map report issued by the WCEC on:** November 2022

#### WCEC Team

- Adrian Edwards, Alison Cooper, Ruth Lewis, Micaela Gal, Jane Greenwell and Helen Freegard involved in drafting the Topline Summary and editing

#### This review should be cited as

REM00043.Wales COVID-19 Evidence Centre. A rapid evidence map of effectiveness of community diagnostic centres. October 2022

#### Disclaimer

The views expressed in this publication are those of the authors, not necessarily Health and Care Research Wales. The WCEC and authors of this work declare that they have no conflict of interest.

### TOPLINE SUMMARY

#### What are Rapid Evidence Maps?

Our Rapid Evidence Maps (REMs) use abbreviated **systematic mapping or scoping review methods** to provide a description of the nature, characteristics, and volume of the available evidence for a particular policy domain or research question. They are mainly based on the assessment of abstracts and incorporate an a priori protocol, systematic search, screening, and minimal data extraction. They may sometimes include critical appraisal, but **no evidence synthesis is conducted.** Priority is given, where feasible, to studies representing robust evidence synthesis. They are designed and used primarily **to identify a substantial focus for a rapid review, and key research gaps in the evidence-base**. (*N.B. Evidence maps are not suitable to support evidence-informed policy development, as they do not include a synthesis of the results*.)

**This report is linked to a subsequent focused rapid review** published as: RR00043_Wales COVID-19 Evidence Centre. What is the effectiveness of community diagnostic centres: a rapid review. November 2022.

#### Who is this summary for?

Welsh Government Technical Advisory Cell (TAC)

#### Background / Aim of Rapid Evidence Map

The COVID-19 pandemic has resulted in increased demand and delays to diagnostic services. **Community diagnostic centres** (which are generally referred to as Regional Diagnostic Hubs in Wales) aim to reduce this backlog and the waiting times for patients by **providing a broad range of elective diagnostic services in the community, away from acute hospital facilities**. As diagnostic services account for over 85% of clinical pathways and cost the NHS over £6 billion a year (NHS 2022), community diagnostic centres across a broader range of diagnostic services may be an effective, efficient, and cost-effective introduction to the UK health sector.

A preliminary review of the literature identified a large volume of primary studies looking at a broad range of outcomes in relation to different types of diagnostic centres. After discussion with the stakeholders, it was agreed that an evidence map would be useful to understand what evidence exists for the effectiveness of community diagnostic centres, enable stakeholders to identify a focus for a rapid review, and highlight the gaps in evidence base. **This Rapid Evidence Map aimed to identify, describe and map the available evidence on the effectiveness of diagnostic centres**. For the purpose of the report, we use the same ‘community diagnostic centre’ descriptor to incorporate the **variety of descriptors** used for these services. However, the different names and definitions provided within individual studies are also outlined and discussed. Due to the **variation in the location** of community diagnostic centres, **studies were not assessed for inclusion according to their location** (i.e., in the community, primary care, or hospital setting), instead, they were **assessed according to the services they provide, who they provide them for, and their accessibility to primary care / community health care services**.

### Key Findings

50 primary studies were identified (sample sizes ranged from nine to 62,333 participants).

*Summary of the evidence base*

- There were 27 comparative studies and 23 non-comparative studies. A **wide range of study designs were used**, including RCTs (n=2), modelling study, quasi-experimental studies, descriptive audits, and evaluation studies looking at a single population as they passed through the diagnostic centre.
- Studies were conducted in a **range of countries including the UK (n=21),** Spain (n=17), Canada (n=8), Australia (n=2), Denmark (n=1) and The Netherlands (n=1). Of the 21 studies conducted in the UK, three were from Wales (one was focussed on an autism diagnostic centre and two were cancer specific).
- **30 studies were specific to cancer diagnosis**, whilst the remaining 20 studies focused on diagnosis associated with: anaemia (n=1), autism (n=1), cerebral palsy (n=1), intellectual disability (n=1), multiple sclerosis (n=1), respiratory conditions (n=1), shoulder pain (n=1) and unexplained fever (n=1). Eleven studies reported information on multi-condition diagnostic centres, rather than a specific condition and one study did not report a diagnosis of interest.
- The **majority of studies were conducted within hospital settings** (n=45). Two studies evaluated diagnostic centres within a community setting. Three studies did not report the location.
- The diagnostic centres offered a **wide range of diagnostic tests and incorporated different staff and facilities**.
- Participants were **mainly referred by GPs (n=23), primary care centres (n=21)** and emergency departments (n=18). However, referrals were also made from outpatient clinics located within the same hospital as the diagnostic centre (n=8), medical specialists (n=5), specialist outpatient clinics (n=3), other settings (the definition of this was not described) (n=3), inpatient wards (n=1), and community specialists (n=1). Four studies did not clearly report where the referrals originated.
- **113 different outcomes were reported** covering: patient data and referral outcomes (n=47); clinical outcomes (n=45); performance outcomes (n=43); economic outcomes (n=22) and patient and physician-reported outcomes (n=21).

#### Recency of the evidence base

- Studies were published between 1995 and 2021, with the data collected between 1993 and 2019

#### Best quality evidence

- **Two RCTs** (Harcourt et al 1998, Dey at al 2002) were identified, both conducted in the UK and were specific to the diagnosis of breast cancer. Both made comparisons between a one-stop clinic and conventional clinic arrangements.

### Implications for a Rapid Review

The findings of the REM were used to select a substantive focus for a subsequent rapid review and the different options discussed at a stakeholder meeting (held on the 13^th^ September 2022). A decision was made to focus on community diagnostic centres that can be accessed by primary care teams, and the evidence relating to any condition should be considered, not just cancer. It was decided that the primary outcomes should align with the need to evaluate whether community diagnostic centres can increase capacity for diagnostics and reduce pressure on secondary care, as well as ensure equity in uptake or access. Economic outcomes were also considered pertinent. Finally, the review should be limited to comparative studies, prioritising evidence from studies using more robust study designs.

## 1. BACKGROUND

### 1.1 Who is this Rapid Evidence Map for?

This Rapid Evidence Map was conducted as part of the Wales COVID-19 Evidence Centre Work Programme. The above question was suggested by the Welsh Government Technical Advisory Cell (TAC).

### 1.2 Purpose of this review

The COVID-19 pandemic has had a direct impact on the number of patients awaiting diagnostic services due to the prioritisation of urgent treatments and the suspension of non-urgent appointments in March 2020 (Welsh Government 2021). The backlog brought about by the pandemic has resulted in an increased demand for diagnostic services, which in turn has resulted in increased waiting times for diagnostics and treatment. In Wales, evidence has shown that the number of patients waiting longer than the target of eight weeks for diagnostics increased by 41.2% between March 2020 and April 2022 (Welsh Parliament 2022). In order to reduce the backlog and delays to diagnostic services, a report of the Independent Review of Diagnostic Services for NHS England recommended the creation of community diagnostic centres (National Health Service England 2020). Community diagnostic centres aim to reduce this backlog and the waiting times for patients by providing a broad range of elective diagnostic services in the community, away from acute hospital facilities. In Wales, community diagnostic centres are generally referred to as **Regional Diagnostic Hubs.**

In England, community diagnostic centres were first launched in 2021 (Department of Health and Social Care 2021), and now over 90 community diagnostic centres have been opened with plans to open up to 160 centres by 2025 (National Health Service England 2022). In Wales, a plan to create a network of community diagnostic centres has also been outlined by the Welsh Government (Welsh Government 2022). As diagnostic services account for over 85% of clinical pathways and cost the NHS over £6 billion a year (National Health Service 2022), community diagnostic centres across a broader range of diagnostic services may be an effective, efficient, and cost-effective introduction to the UK health sector. These services will ensure timely diagnosis and reduced waiting times and would make sure people receive the right treatment or get referred to the right specialists.

Our initial investigation into this topic identified a large number of primary studies looking at a broad range of outcomes in relation to diagnostic centres. After discussion with stakeholders, it was agreed a rapid evidence map (REM) would be useful to understand the available evidence.

This REM seeks to understand what available evidence exists for the effectiveness of community diagnostic centres. This will be used to identify a substantive focus for a subsequent Rapid Review and highlight gaps in the evidence base.

### 1.3 Definition of diagnostic centres

For the purpose of this REM, community diagnostic centres are defined as health services aimed at improving population health outcomes by providing quicker and easily accessible diagnostic services in the community, which are accessible to primary care practitioners/services, thereby relieving pressure on secondary care services.

In Wales, community diagnostic centres are generally referred to as Regional Diagnostic Hubs to avoid confusion with the descriptors or acronyms used for other similar services. Diagnostic centres are also described within the international literature using a variety of terms and definitions. For the purposes of this REM, we will therefore use the same descriptor ‘community diagnostic centres’ to incorporate the range of terms used for these services. However, the different descriptors used within individual studies are also outlined and discussed in this report.

Due to the variation in the location of community diagnostic centres, studies were not assessed for inclusion according to their location (i.e., in the community, primary, secondary, or tertiary care), instead, they were assessed according to the services they provide, who they provide them for, and that they are accessible to primary care practitioners/services.

## 2. FINDINGS

### 2.1 Overview of the available evidence

Our searches identified a total of 50 primary studies which were published between 1995 and 2021. Data from the primary studies were collected between 1993 and 2019 and sample sizes ranged from nine to 62,333 participants.

We have mapped the location of the diagnostic centres, the diagnosis of interest, and referring physicians or services to the centres, see Table 1.

- We identified 30 primary studies specific to cancer diagnosis and 20 reporting other conditions (hereafter known as non-cancer specific). Non-cancer specific diagnostic centres covered a range of conditions including anaemia (n=1), autism (n=1), cerebral palsy (n=1), intellectual disability (n=1), multiple sclerosis (n=1), respiratory conditions (n=1), shoulder pain (n=1) and unexplained fever (n=1). Eleven studies reported information on multi-condition diagnostic centres, rather than a specific condition and one study did not report a diagnosis of interest.
- The majority of studies were conducted within hospital settings (n=45). Two studies evaluated diagnostic centres within a community setting while three studies did not report the setting or were unclear on the location of the diagnostic centre.
- Participants were most commonly referred to the diagnostic centres by GPs (n=23), primary care centres (n=21) and emergency departments (n=18). However, referrals were also made from outpatient clinics located within the same hospital as the diagnostic centre (n=8), medical specialists (n=5), specialist outpatient clinics (n=3), other settings (the definition of this was not described) (n=3), inpatient wards (n=1), and community specialists (n=1). Four studies did not clearly report where the referrals originated.
- Although three of the primary studies examined multiple diagnostic centres located within different hospitals, only 29 individual diagnostic centres were reported on across the 50 included studies in total.
- The diagnostic centres described within the included studies offered a range of diagnostic tests which can be seen in Appendix 1 (Diagnostic centre characteristics table), along with details about the location and setting of the diagnostic centre, staff and facilities, diagnostic tools, conditions, referrals, and referral criteria.

**Table 1.**
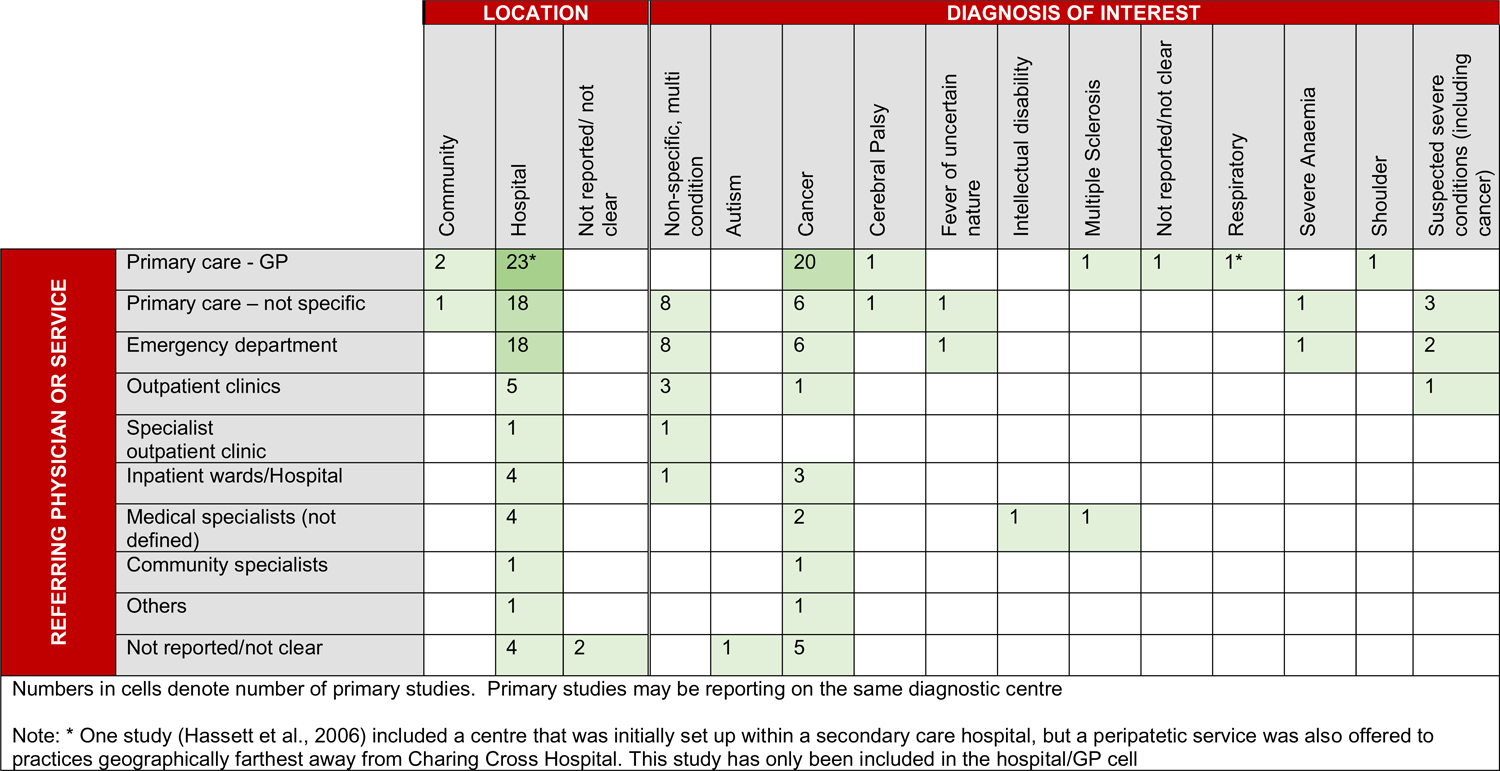
Number of studies per location, diagnosis of interest and referring physician or service

### 2.2 Methodology of studies identified

We identified a wide range of study designs among our included studies, including descriptive audits and evaluation studies looking at a single population as they passed through the diagnostic centre. We also identified two RCTs, one modelling study, and some quasi-experimental studies. Some studies compared populations attending a new diagnostic centre with patients who had been diagnosed prior to the opening of a diagnostic centre (this was often classed as usual care). Some studies also compared two populations attending different diagnostic centres. Many studies utilised convenience sampling methods, and some conducted telephone interviews with patients and referring physicians three months or more after the patient had visited the diagnostic centres.

Due to the inconsistent and often poorly reported methods in most of the included studies, it has not been possible to identify the methodology used in the included studies in the given timeframe stipulated for this REM. In order to make inferences about the impact or effectiveness of diagnostic centres, it is important to identify the study methodology used so that only those studies using appropriate methodology are selected. We trialled an algorithm produced by Leatherdale (2019) to classify types of ‘natural experiment’ study designs, but this was not successful in identifying the study designs of most of our included studies. Any further analysis undertaken on the effectiveness of diagnostic centres will need to identify an appropriate tool to ensure only studies with an appropriate methodology are included.

### 2.3 Country of origin of included studies

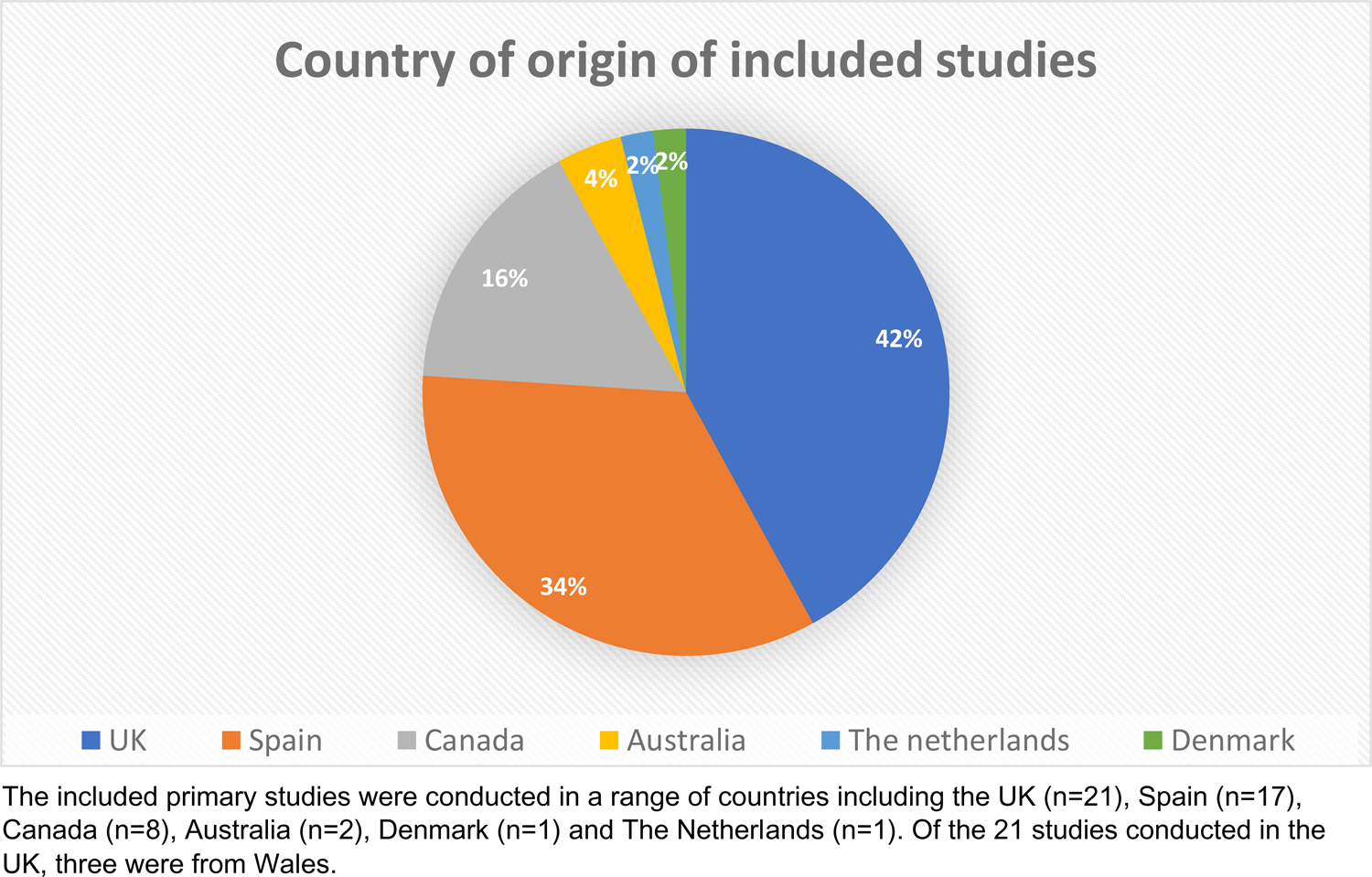

The 50 primary studies included in this REM reported findings from 29 individual diagnostic centres. This included 17 diagnostic centres reported across the 21 UK studies, four diagnostic centres across the 17 Spanish studies, and four diagnostic centres across the eight Canadian studies. The two studies conducted in Australia reported on two separate diagnostic centres and only one diagnostic centre was included in the singe study from Denmark, as with the single study conducted in the Netherlands. As many of the studies evaluated and collected data on the same diagnostic centre and around the same time period it is possible that the same data were reported across multiple studies. Appendix 2 includes details on the data collection period, study design, population, outcomes, study comparison details and information on the potential risk of data being reported across multiple studies.

It is likely that the large number of Spanish studies identified utilised the same datasets. However, this was not possible to ascertain within our given timeframe because these studies included different (but often overlapping) data collection dates, various populations and often reported different outcomes. As a result, it was decided to treat all Spanish studies as individual studies within the maps.

### 2.4 Outcomes measured

Across the 50 included primary studies a total of 113 outcomes were reported, these outcomes were grouped into five categories:

- Patient demographics and referral outcomes
- Clinical outcomes
- Performance outcomes
- Economic outcomes and
- Patient and physician-reported outcomes.

Outcomes have been mapped against the location of the diagnostic centre and the referring physician or service in Table 2. Outcomes have also been mapped against the diagnosis of interest in Table 3. Detailed maps of the outcomes reported by each individual study can be seen in Appendix 3.

**Table 2:** Number of studies by location, referring physician or service and outcomes

|  |  | LOCATION |  |  | REFERRING PHYSICIAN OR SERVICE |  |  |  |  |  |  |  |  |  |
| --- | --- | --- | --- | --- | --- | --- | --- | --- | --- | --- | --- | --- | --- | --- |
|  |  | Community (n=2) | Hospital (n=45) | Not reported/ not clear (n=3) | Primary care – GP (n=23) | Primary care – not specific (n=21) | Emergency department (n=18) | Outpatients clinic (n=8) | Specialist outpatient clinic (n=3) | Inpatient ward/hospital (n=1) | Medical specialists (not defined) (n=5) | Community Specialist (n=1) | Others (n=3) | Not reported/not clear (n=4) |
| OUTCOMES REPORTED | Patient demographics and referral | 2 | 44 | 2 | 23 | 21 | 18 | 5 | 2 | 1 | 5 | 1 | 3 | 3 |
|  | Performance | 2 | 39 | 1 | 21 | 19 | 18 | 5 | 2 | 1 | 5 | 1 | 3 | 3 |
|  | Clinical | 2 | 40 | 1 | 20 | 16 | 18 | 6 | 3 | 1 | 4 | 1 | 3 | 3 |
|  | Economic | 1 | 20 |  | 15 | 14 | 12 | 4 |  |  | 3 |  | 3 | 1 |
|  | Patient and physician-reported | 2 | 17 | 2 | 15 | 7 | 6 | 2 | 1 |  | 4 |  | 2 | 2 |

**Table 3:** Number of studies by diagnosis of interest and outcomes

|  |  | DIAGNOSIS OF INTEREST |  |  |  |  |  |  |  |  |  |  |  |
| --- | --- | --- | --- | --- | --- | --- | --- | --- | --- | --- | --- | --- | --- |
| OUTCOMES REPORTED |  | Non-specific Multi-condition (n=8) | Autism (n=1) | Cancer (n=30) | Cerebral Palsy (n=1) | Fever of uncertain nature (n=1) | Intellectual disability (n=1) | Multiple Sclerosis (n=1) | Not reported/not clear (n=1) | Respiratory (n=1) | Severe Anaemia (n=1) | Shoulder (n=1) | Suspected severe conditions (including cancer) (n=3) |
|  | Patient demographics and referral | 8 |  | 28 | 1 | 1 | 1 | 1 | 1 | 1 | 1 | 1 | 3 |
|  | Performance | 8 |  | 24 | 1 | 1 | 1 | 1 | 1 | 1 | 1 |  | 3 |
|  | Clinical | 8 |  | 25 | 1 | 1 | 1 | 1 | 1 | 1 | 1 | 1 | 2 |
|  | Economic | 4 |  | 9 |  | 1 | 1 | 1 | 1 |  | 1 | 1 | 2 |
|  | Patient and physician-reported | 2 | 1 | 11 | 1 |  | 1 |  | 1 | 1 | 1 |  | 1 |

**Table 4.** Eligibility criteria

|  | **Inclusion criteria** | **Exclusion criteria** |
| --- | --- | --- |
| **Participants** | Symptomatic patients, all conditions |  |
| **Settings** | Diagnostic centres | Exclude screening programmes, or where there is treatment undertaken but no diagnostics. |
| **Intervention / exposure** | Diagnostic centres/units/hubs and clinics |  |
| **Comparison** | Usual care |  |
| **Outcomes** | All reported outcomes<br>e.g.<br>Mortality<br>Clinical outcomes<br>Patient reported outcomes<br>Health care professional reported outcomes<br>Resources<br>Costs<br>Process outcomes<br>Workforce |  |
| **Study design** | Any design |  |
| **Countries** | All countries |  |
| **Language of publication** | Studies published in English | Any study not published in English |
| **Publication date** | No date limits set |  |
| **Publication type** | Published and preprint primary literature | All publication types other than primary literature |

## 3. DISCUSSION

### 3.1 Summary of the findings

A total of 50 primary studies were identified and reported a range of outcomes exploring the effectiveness of community diagnostic centres.

- A total of 47 studies reported patient demographics and referral outcomes, 45 studies reported clinical outcomes, 43 studies reported performance outcomes, 22 studies reported economic outcomes and 21 studies reported patient and physician-reported outcomes.
- The least common outcome group reported across all diagnosis of interest was patient and physician-reported outcomes. A higher number of cancer specific studies reported on each outcome group compared to the non-cancer specific studies, apart from economic outcomes, which was reported by 12 non-cancer specific studies and only nine cancer-specific studies.
- Only one study was conducted on a diagnostic centre for autism (Kerrell 2001) and this study only reported patient and physician-reported outcomes. However, all other diagnoses of interest reported outcomes in at least three of the outcome categories.
- The two RCTs (Harcourt et al 1998, Dey at al 2002) were both conducted in the UK and were specific to the diagnosis of breast cancer. Both made comparisons between a one-stop clinic and conventional clinic arrangements.
- Of the three studies conducted in Wales, one was focussed on an autism diagnostic centre (Kerrel 2001) and two were cancer specific (Sewell et al 2020, Vasilakis et al 2021).

### 3.2 Limitations of the available evidence/evidence gaps

The maps have highlighted several evidence gaps. As the majority of studies identified were specific to cancer diagnostic centres (n=30), further research is needed to evaluate diagnostic centres that are specific to a range of other conditions.

Only two studies described a diagnostic centre as being set within the community (rather than hospital-based) and as such further evidence is needed in order to understand how situating diagnostic centres in locations other than hospitals can be of benefit in terms of who accesses them and what effect this can have on waiting times.

It is possible that many of the primary studies identified are not methodologically robust enough to make inferences about the effectiveness of diagnostic centres. However, this has not been investigated in detail, but what is clear is that there was a wide range of methodologies applied to the included primary studies.

Although many of the included studies reported on the same diagnostic centre, the data collection dates typically varied, however many of the Spanish studies appeared to use the same datasets. This could introduce errors to review findings. Any further rapid review will need to take this into consideration and take steps to avoid potential double counting of the same data across multiple studies.

We identified two RCTs, however, the majority of studies included were descriptive in design and therefore not suitable for evaluating the effectiveness of interventions. Any further work conducted after this map will need to take this into account if the focus of the question is about effectiveness of diagnostic centres.

### 3.3 Strengths and limitations of this Rapid Evidence Map

The primary studies included in this REM were identified through an extensive search of electronic databases, trial registries, grey literature, as well as consultation of content experts in the field. Despite making every effort to capture all relevant publications and reduce the risk of bias, it is possible that additional eligible publications may have been missed or we may have introduced some biases to this REM. We have highlighted this where possible, for example in the investigation into the potential risk of multiple studies reporting the same data.

As we did not set date or country limits, and the data collection dates of the included primary studies are wide ranging, it is possible that the diagnostic centres we have included here may not be the same as the proposed diagnostic centres within Wales. It is also possible that as many of the diagnostic centres included were from other countries where the healthcare system is different to that of the UK, the results may not be generalisable to the UK. As this is a REM, we did not conduct any data synthesis or quality appraisal of included evidence, therefore we cannot report the quality of the studies or overall effectiveness of community diagnostic centres.

### 3.4 Implications and next steps

A stakeholder meeting was held on the 13^th^ September 2022 where the findings of the REM were presented. Given the available evidence, it was decided to conduct a Rapid Review (RR) of the literature focussing on the effectiveness of community diagnostic centres that accepted referrals from a primary care setting. Due to the often descriptive nature of many of the included studies it was also decided that the RR would only include comparative studies prioritising evidence from studies using more robust study designs.

It was agreed with stakeholders that the primary outcomes should align with the need to evaluate whether community diagnostic centres can increase capacity for diagnostics and reduce pressure on secondary care, as well as ensure equity in uptake or access. Economic outcomes will also be considered as a secondary outcome measure.

## Abbreviations

Acronym: Full Description

CDC: Complex Diagnostic Clinic

CMV: Cytomegalovirus

COVID-19: Coronavirus Disease 2019

CT: Computed Tomography

DDC: Demyelinating Disease Diagnostic Clinic

EBV: Epstein–Barr virus

ED: Emergency Department

ERCP: Endoscopic Retrograde Cholangiopancreatography

FNPA: Fine-Needle Puncture Aspiration

FUN: Fever of Uncertain Nature

GNC: General Neurology Clinic

GP: General Practitioner

HIV: Human Immunodeficiency Virus

IIU: Inpatient Investigation Unit

MRI: Magnetic Resonance Imaging

NHS: National Health Service

PCC: Primary Care Centres

PET-CT: Positron Emission Tomography-Computed Tomography

PHC: Primary Healthcare

REM: Rapid Evidence Map

RR: Rapid Review

RCT: Randomised Controlled Trial

QDU: Quick Diagnosis Unit

TAC: Technical Advisory Cell

UK: United Kingdom

WCEC: Welsh Covid-19 Evidence Centre

## 5. RAPID EVIDENCE MAP METHODS

### 5.1 Eligibility criteria

We searched for primary sources to answer the review question “What is the available evidence for the effectiveness of community diagnostic centres?”

The following eligibility criteria were used to identify studies for inclusion in the REM:

### 5.2 Literature search

COVID-19 specific and general repositories of evidence reviews noted in our resource list were searched on 6^th^ July 2022 by three reviewers and an updated search was conducted on the 3^rd of^ August 2022. An audit trail of the search process is provided within the resource list (Appendix 4). Searches were limited to English-language publications and included searches for primary studies. References of secondary sources identified during preliminary work were scanned for relevant primary studies and forward and backward citation tracking was also conducted on the secondary sources. This included a mapping exercise and focussed rapid review by Chambers et al (2016) who identified current models of community diagnostic services in the UK and internationally and to identify evidence to support a broader range of diagnostic tests being provided in the community. Although this comprehensive work did not meet our inclusion criteria due to the focus being much broader than diagnostic centres, we draw the reader’s attention to it.

Search concepts and keywords around diagnostic units, centres, hubs, and clinics were utilised. The searches combined free text words and descriptors when available. We deliberately kept our search strategy broad to capture as much evidence on diagnostic centres as possible. Resources searched during the REM are outlined in Appendix 4 and the search strategy used to search Medline is available in Appendix 5.

### 5.3 Study selection process

The searches conducted yielded a total of 4,492 records, 349 additional records were identified through citation tracking of secondary sources resulting in a total of 4,841 records. Records were imported into an Endnote database library and duplicates were removed. After deduplication, a total of 3,653 records remained. The title and abstract of the 3,653 records were screened by one reviewer using Rayyan and if relevant, the full text was also screened using the eligibility criteria from section 5.1. A second reviewer consistency checked all the studies selected for inclusion. If disagreements arose, these were discussed, and a third reviewer was consulted to make a final inclusion decision. After full text screening a total of 50 records met the inclusion criteria for this REM. Section 6.1 outlines this process.

### 5.4 Data extraction and coding/charting

Data extraction was undertaken based on the full text and conducted by one reviewer. Information extracted included: the countries where the studies were conducted, sample sizes, publication, and data collection dates. Details about the diagnostic centres including the location and setting, staff and facilities, diagnostic tools, diagnosis of interest, and referral criteria have been reported in Appendix 1: diagnostic centre characteristics table.

The maps utilised a basic coding structure. With 113 individual outcomes reported across the studies, coding was used to group the outcome measures into five categories. Patient demographics and referral outcomes included individual outcomes such as patient characteristics, referral source and reasons for referral. Performance outcomes included individual outcomes such as time to first visit, the number of visits and time to diagnosis. Clinical outcomes included diagnostic yield, diagnostic tests, and onward referrals. Economic outcomes included cost per patient, direct and indirect costs of running the diagnostic centres and overall cost saving. Lastly, patient and physician-reported outcomes included patient satisfaction, referring physician satisfaction and service quality.

Diagnostic location has been categorised as community, hospital or not clear/not reported. This coding was based on the information provided within the primary studies. Primary studies reporting on diagnostic centres set within hospital settings included a range of secondary and tertiary level hospitals, as well as the term hospital only. As many of the tertiary hospitals were set in Spain and other countries outside of the UK, and the lack of information provided within the primary studies, the evidence team were unable to ascertain if the typology used to describe the diagnostic centres was the same as for the UK. The hospital location includes those within secondary and tertiary hospital settings, it was not possible to differentiate where within the community these diagnostic centres were located, so this broad descriptor was used. Unclear/not reported refers to primary studies where it was not possible to ascertain where the diagnostic centre was located.

Descriptors used for ‘referring physician or service’ and ‘diagnosis of interest’ were derived from the primary studies during data extraction. Referrals were often accepted from multiple locations and as such single studies have been reported in multiple categories. Diagnosis of interest was usually a single condition, but some studies reported multiple conditions being diagnosed in a single diagnostic centre. These have been categorised as non-specific and it should be noted that these often included a cancer diagnosis among other conditions.

## 6. EVIDENCE

### 6.1 Study selection flow chart

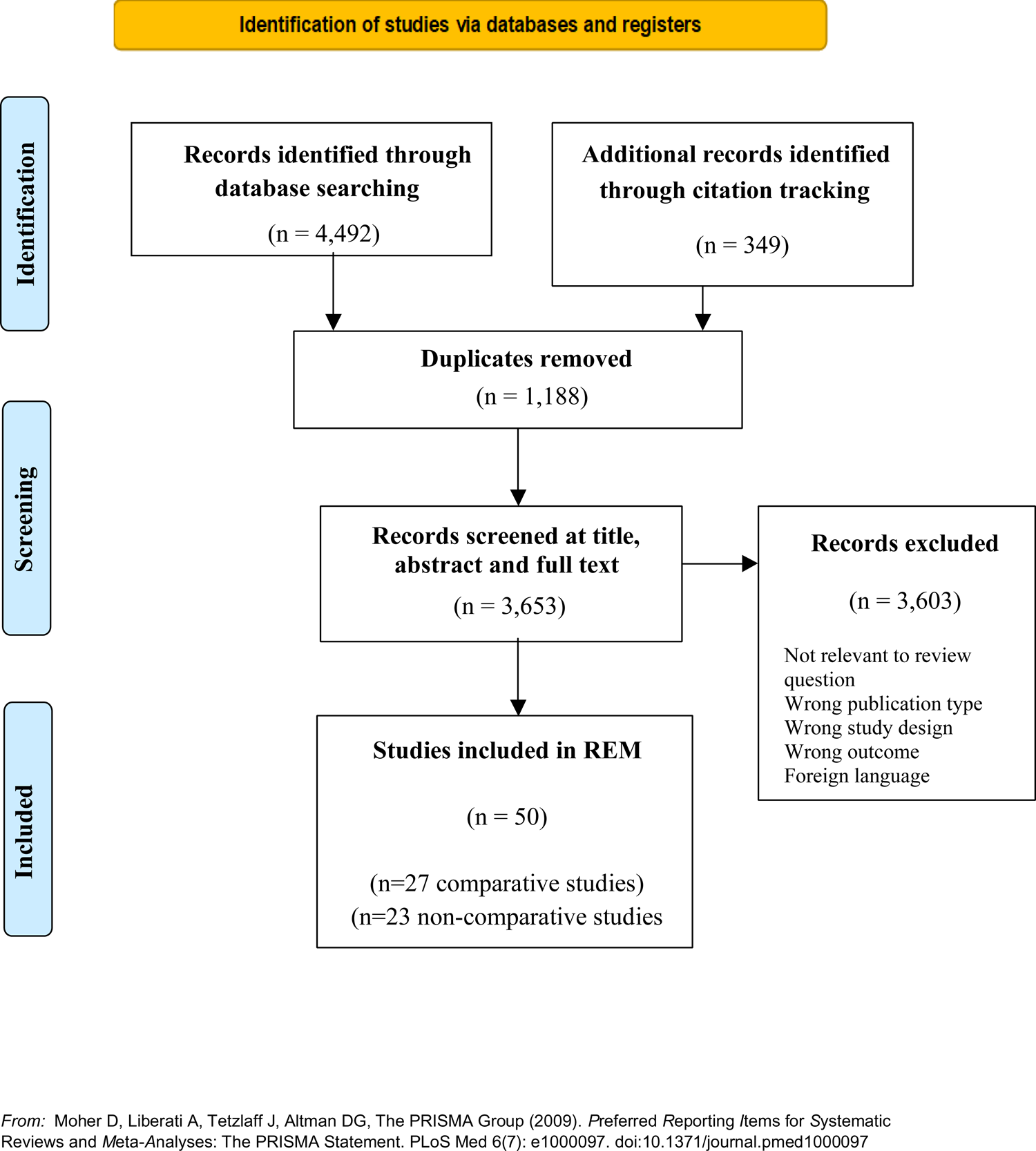

## Data Availability

All data produced in the present study are available upon reasonable request to the authors

## 7.2 Acknowledgements

The authors would like to thank Brendan Collins, Delia Ripley, Jennifer Morgan, Joanna Charles, Leon Wong, Rob Orford and Sally Anstey for their contributions during stakeholder meetings in guiding the focus of the review and interpretation of findings.

## 8. ABOUT THE WALES COVID-19 EVIDENCE CENTRE (WCEC)

The WCEC integrates with worldwide efforts to synthesise and mobilise knowledge from research.

We operate with a core team as part of Health and Care Research Wales, are hosted in the Wales Centre for Primary and Emergency Care Research (PRIME), and are led by Professor Adrian Edwards of Cardiff University.

The core team of the centre works closely with collaborating partners in Health Technology Wales, Wales Centre for Evidence-Based Care, Specialist Unit for Review Evidence centre, SAIL Databank, Bangor Institute for Health & Medical Research/ Health and Care Economics Cymru, and the Public Health Wales Observatory.

Together we aim to provide around 50 reviews per year, answering the priority questions for policy and practice in Wales as we meet the demands of the pandemic and its impacts.

### Director

Professor Adrian Edwards

### Website

https://healthandcareresearchwales.org/about-research-community/wales-covid-19-evidence-centre

## 9. APPENDIX

**APPENDIX 1.**
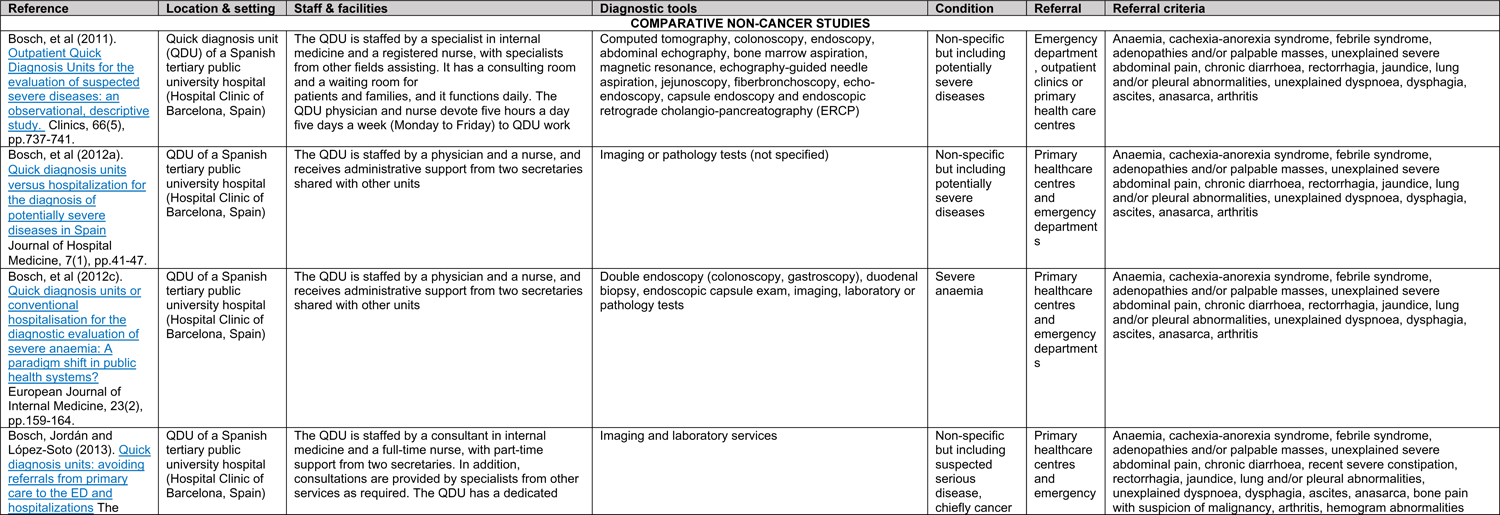
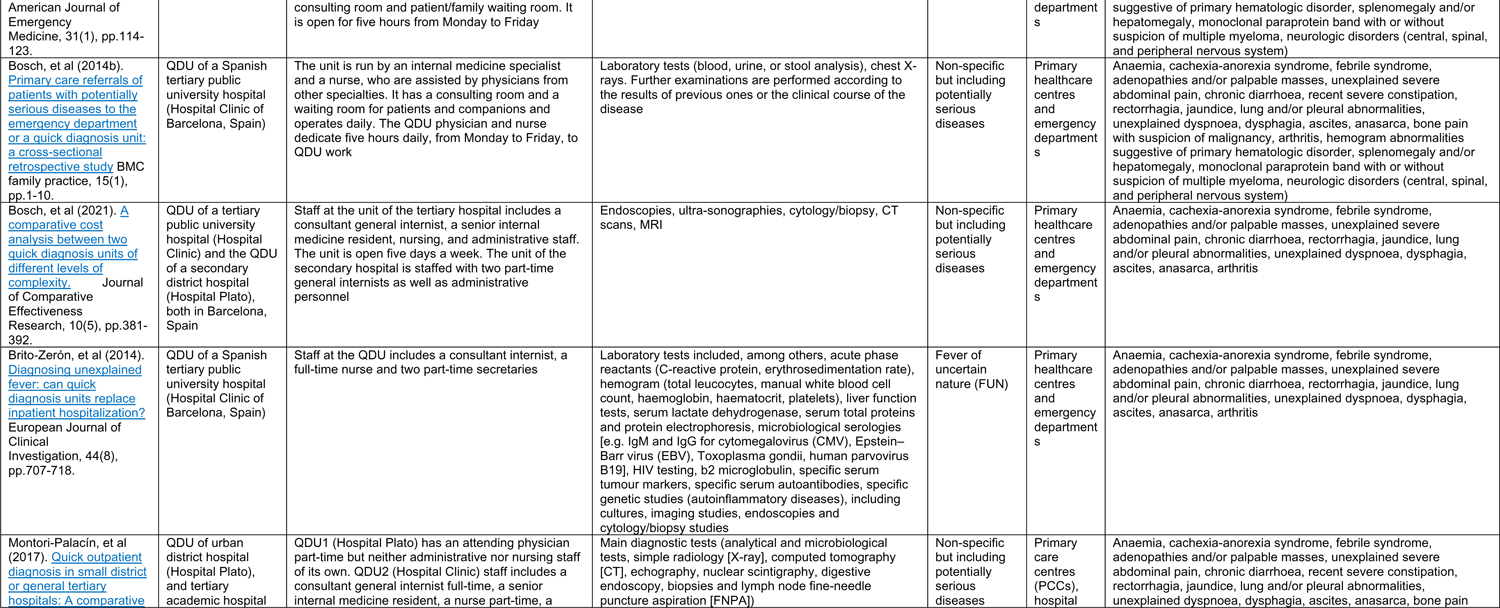
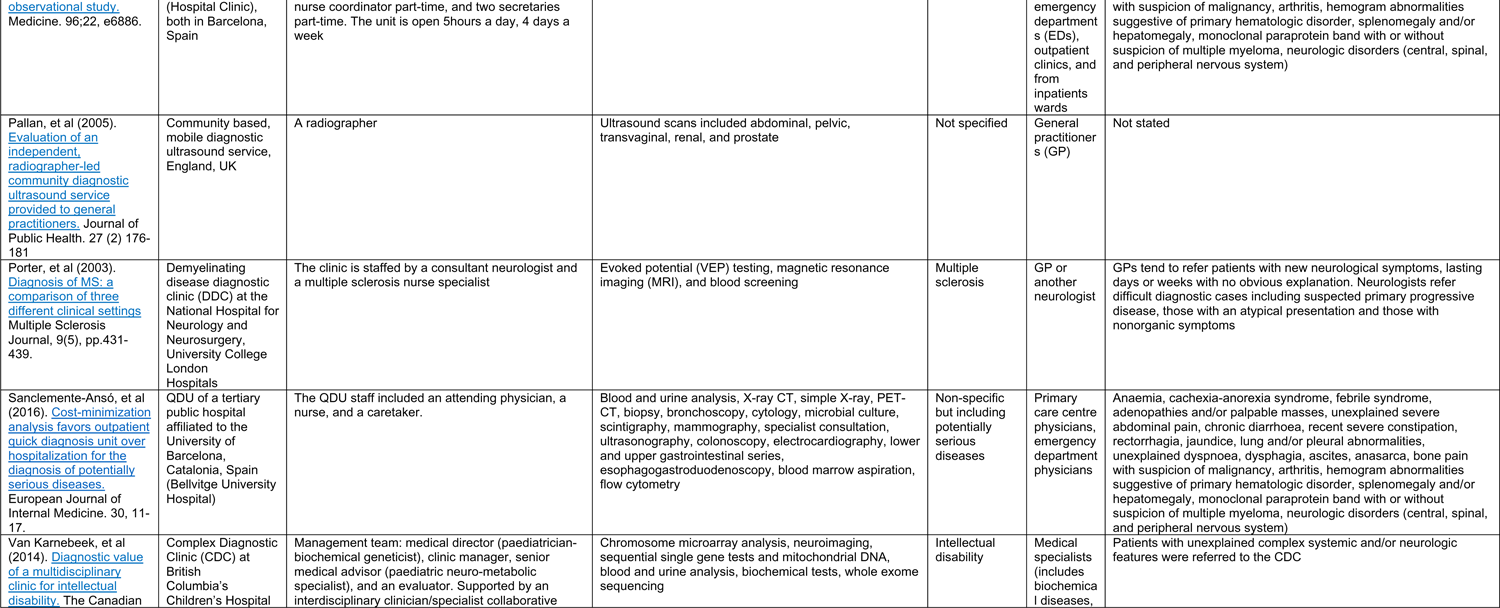
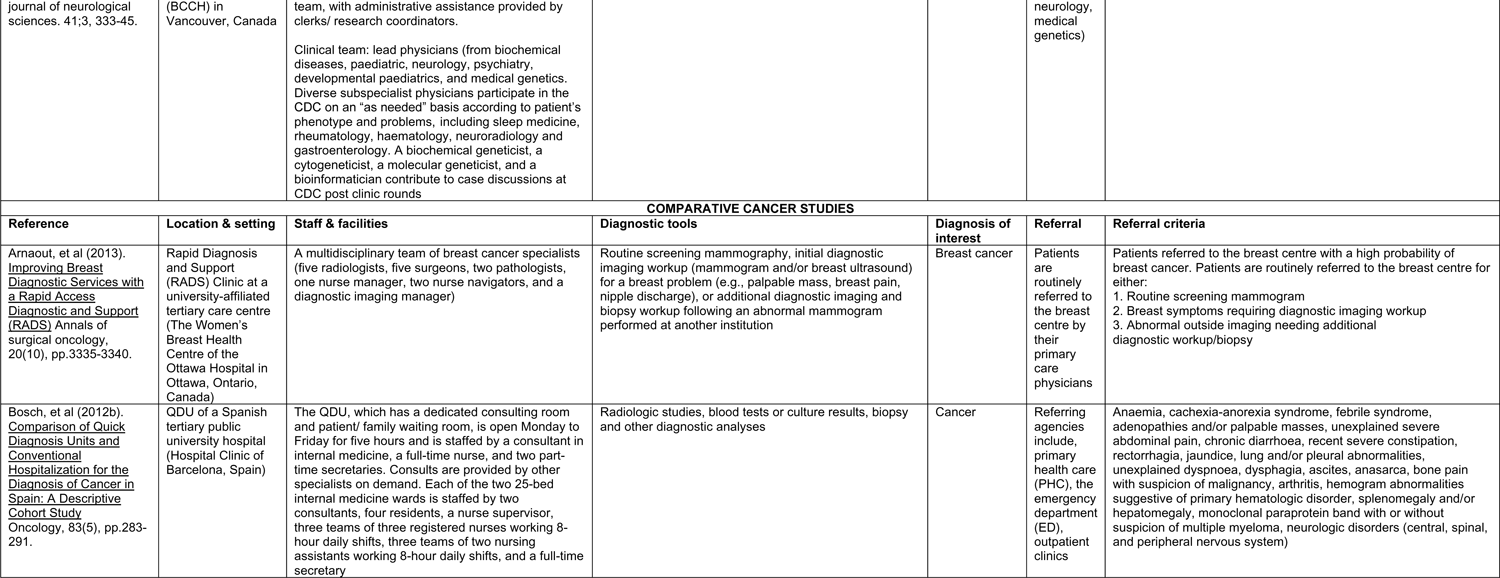
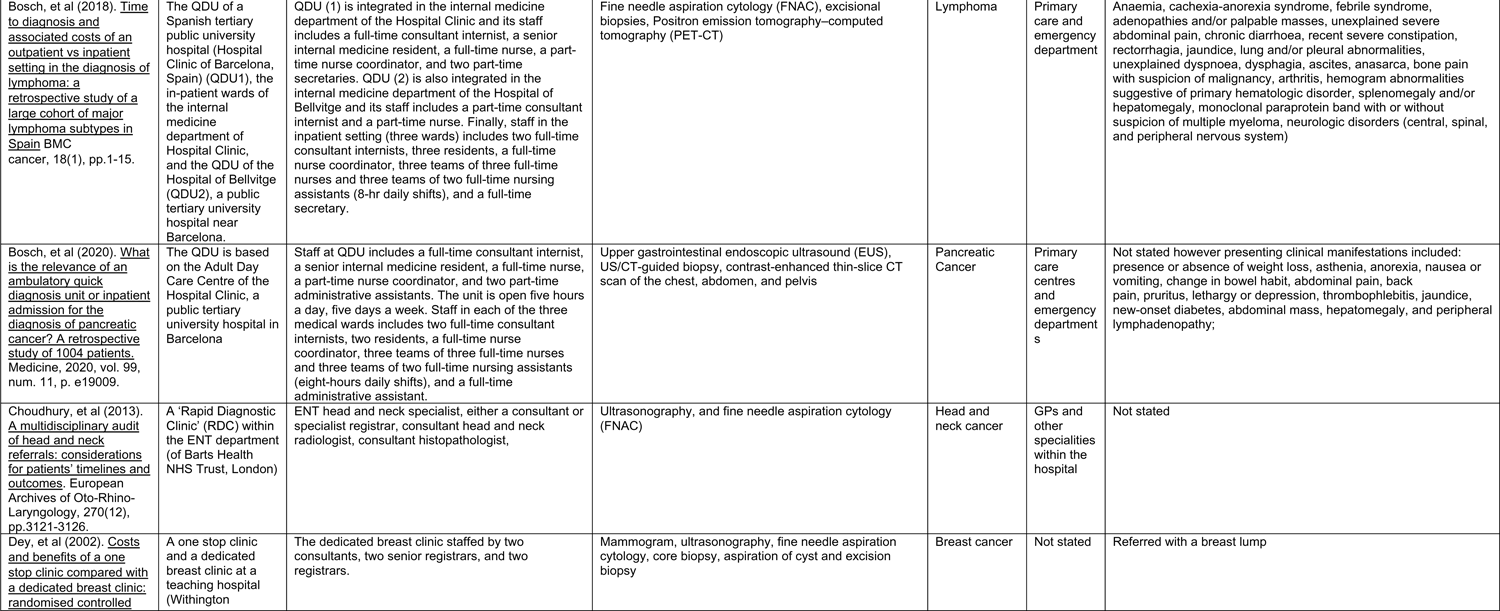
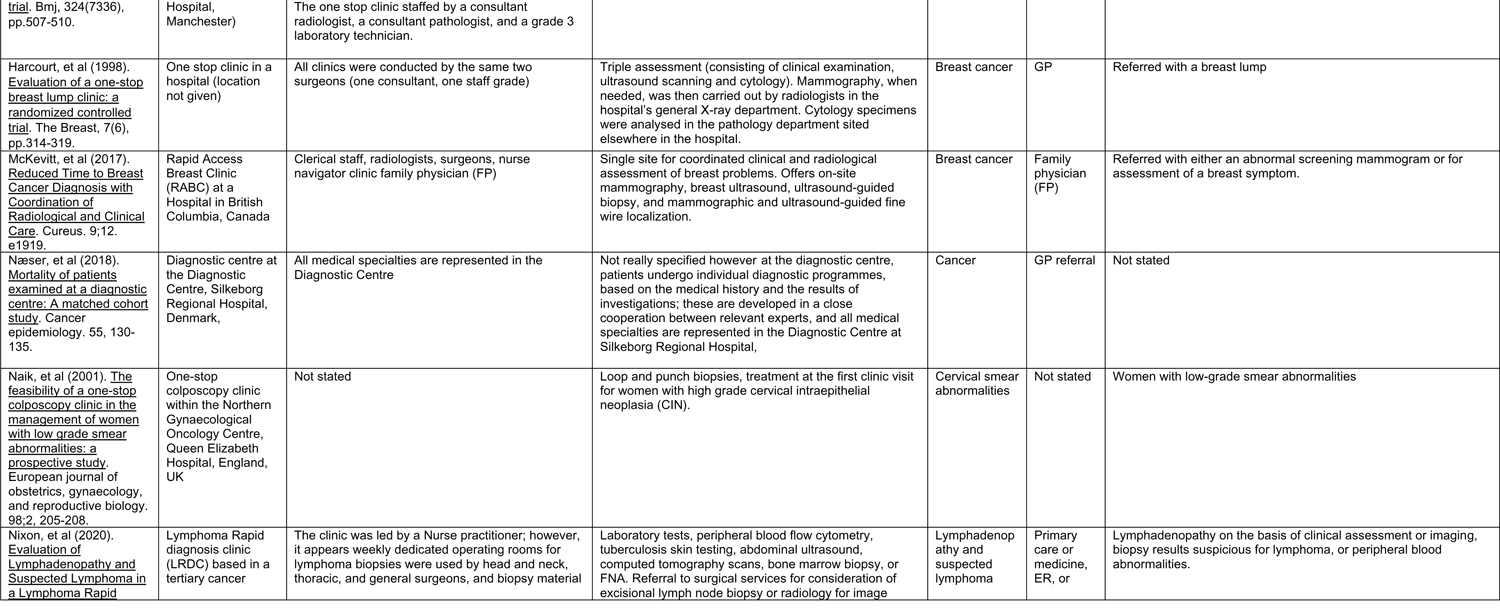
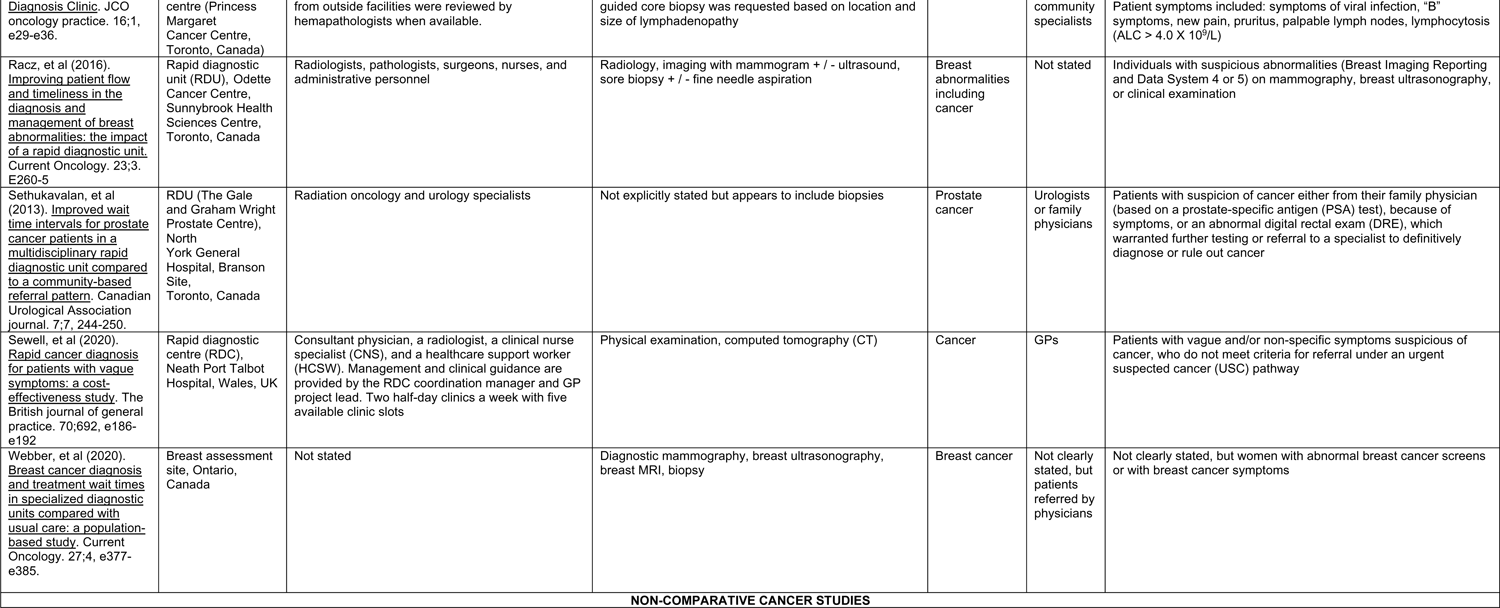
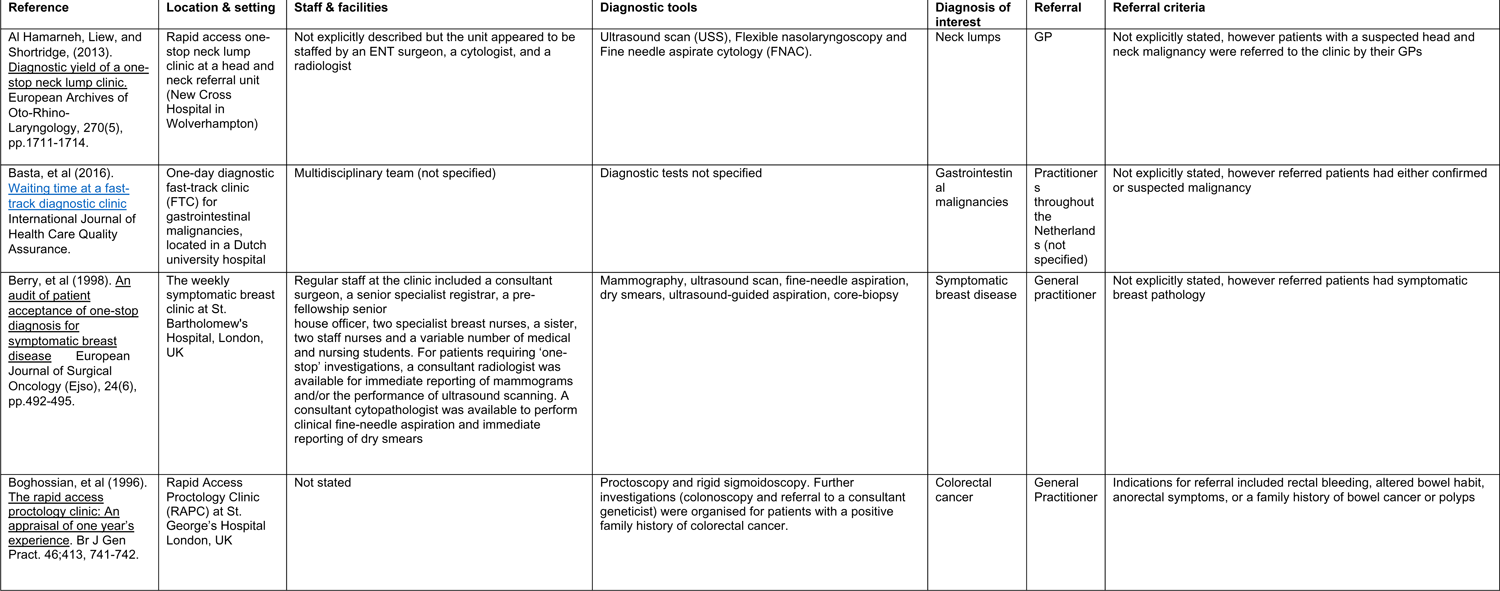
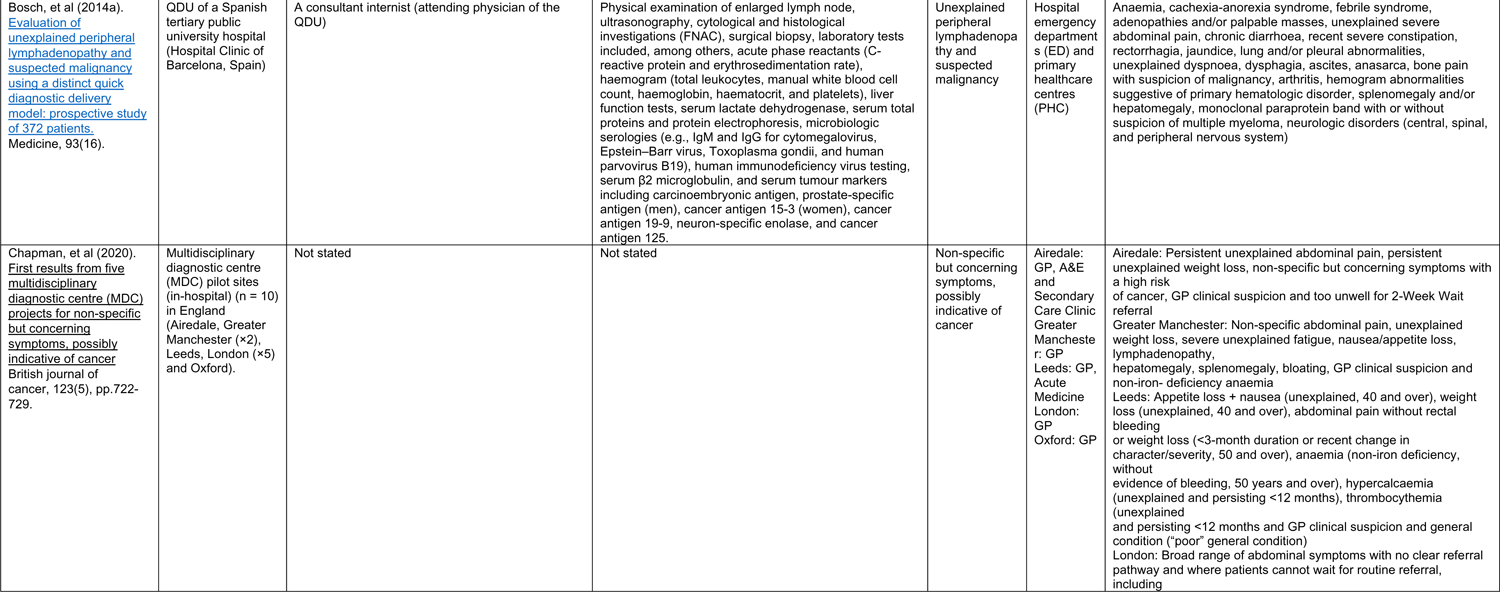
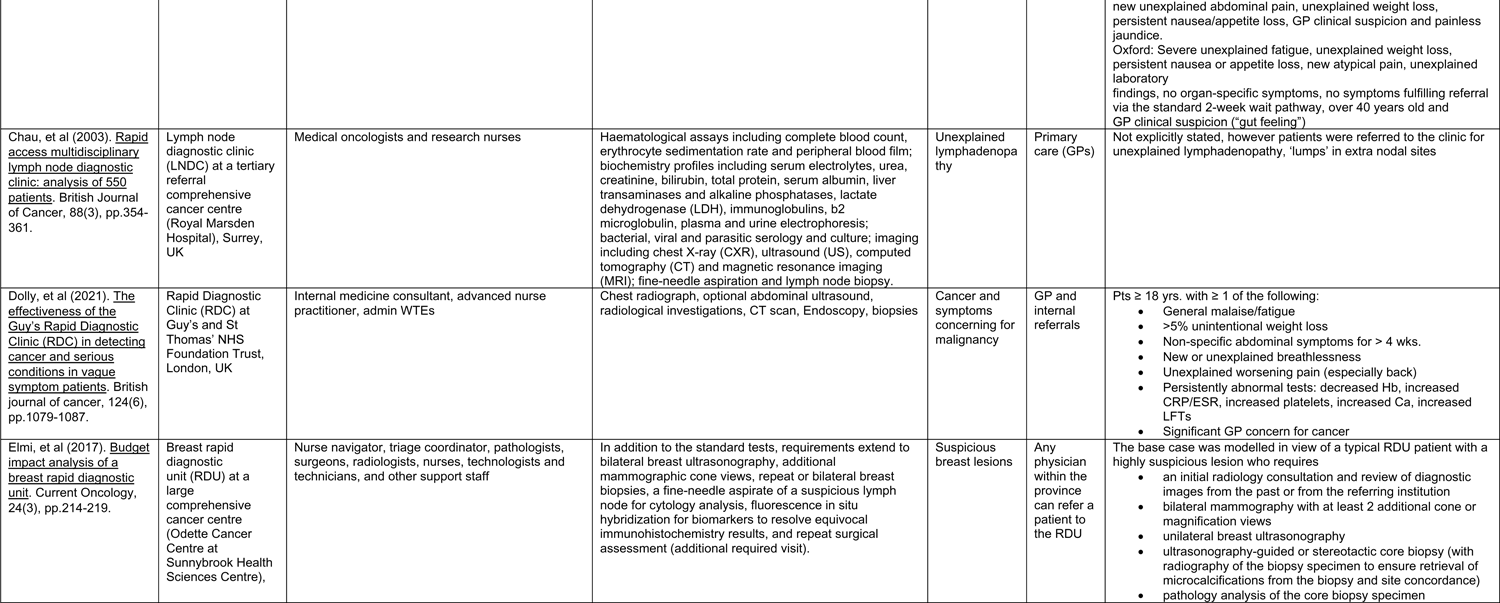
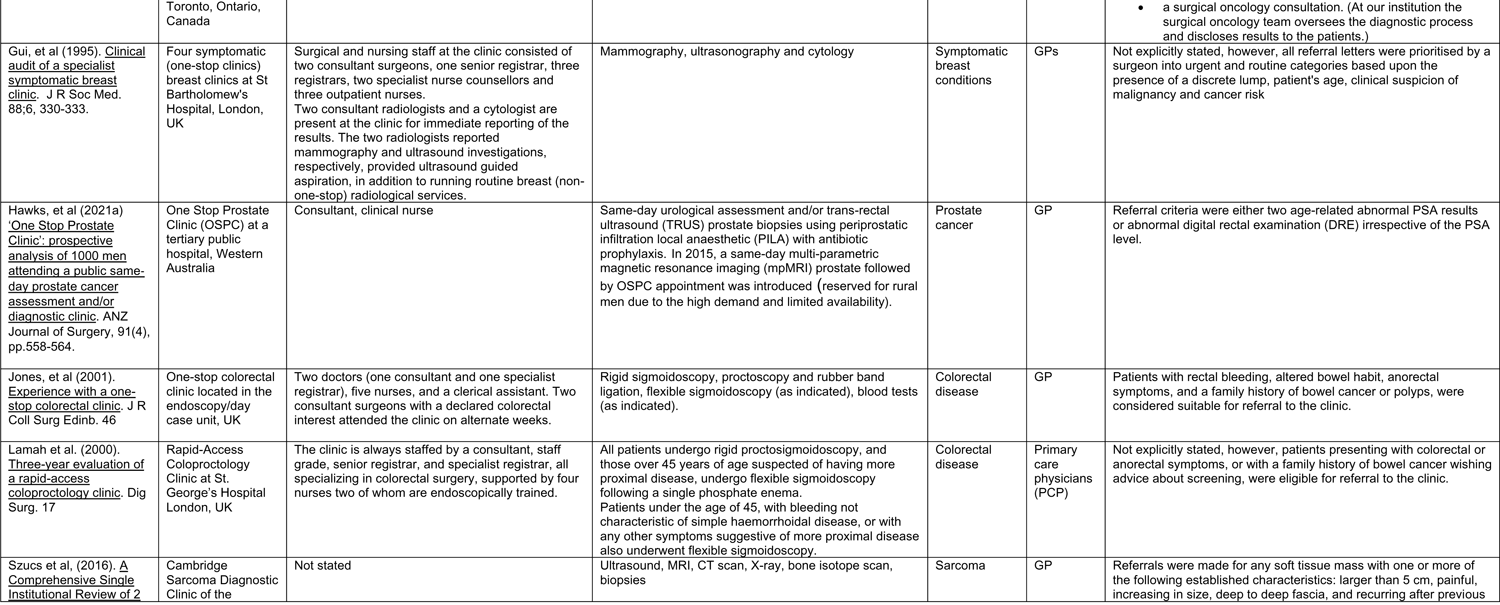
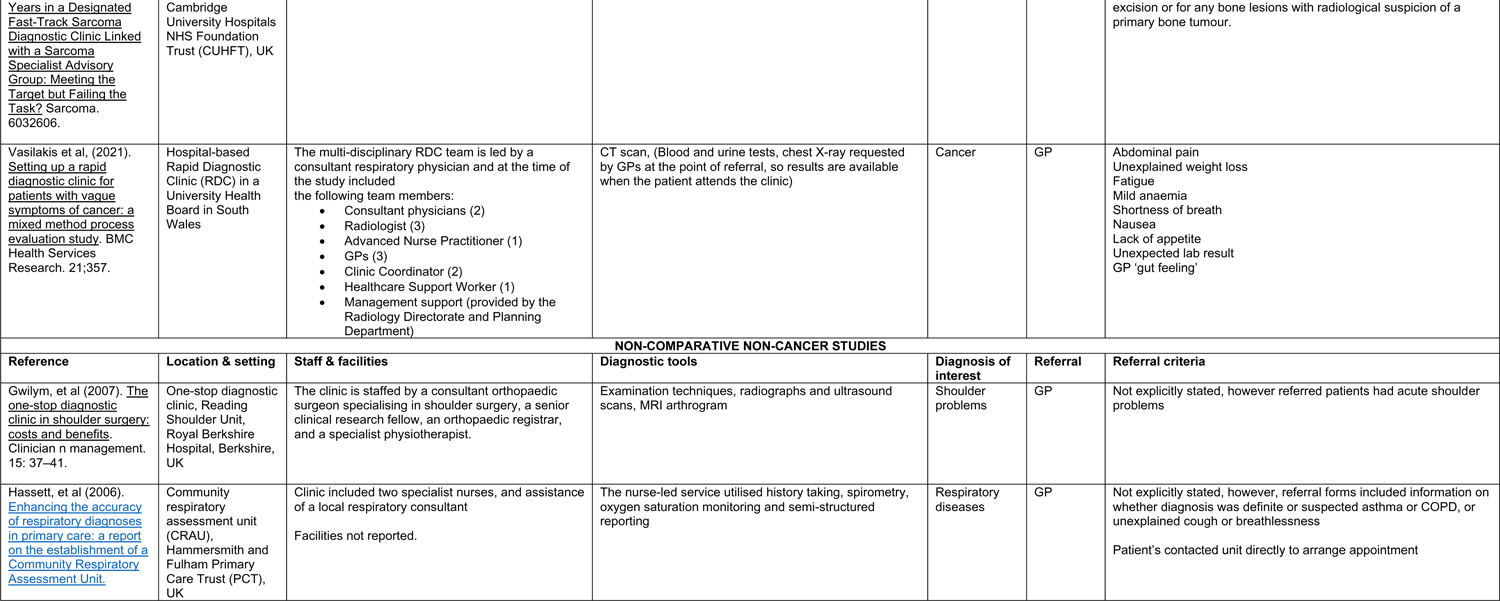
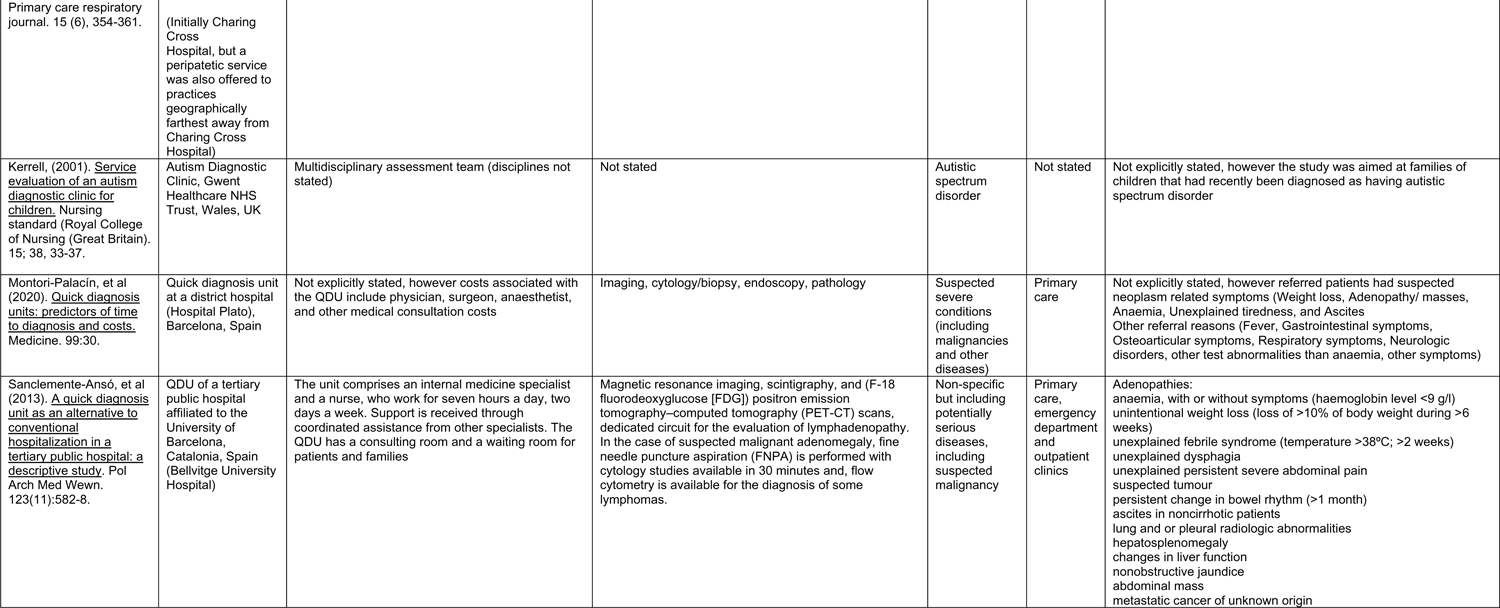
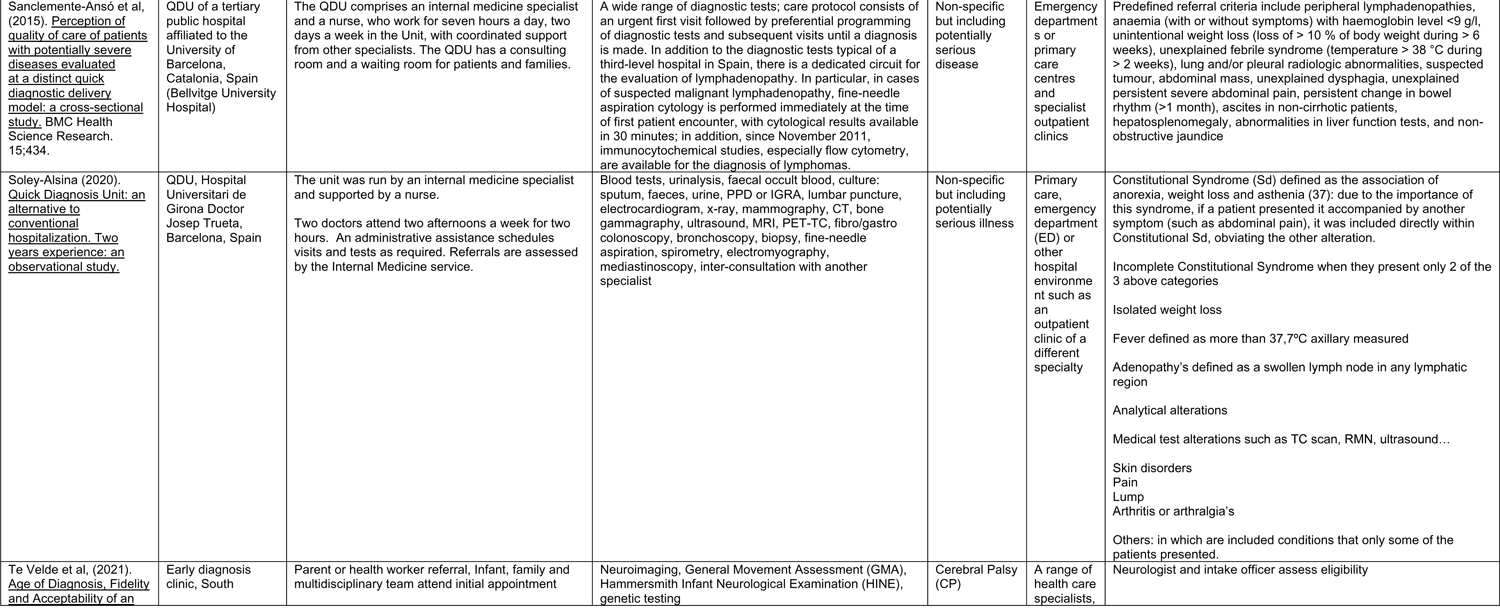
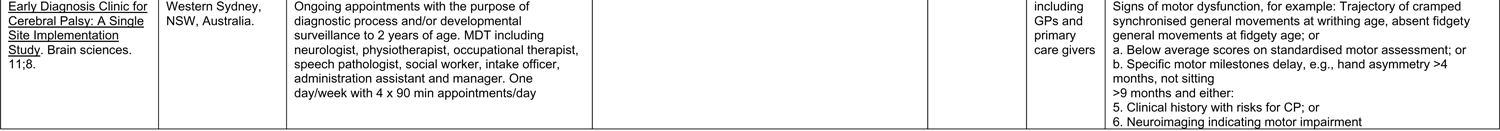
Diagnostic centre characteristics table

**APPENDIX 2.**
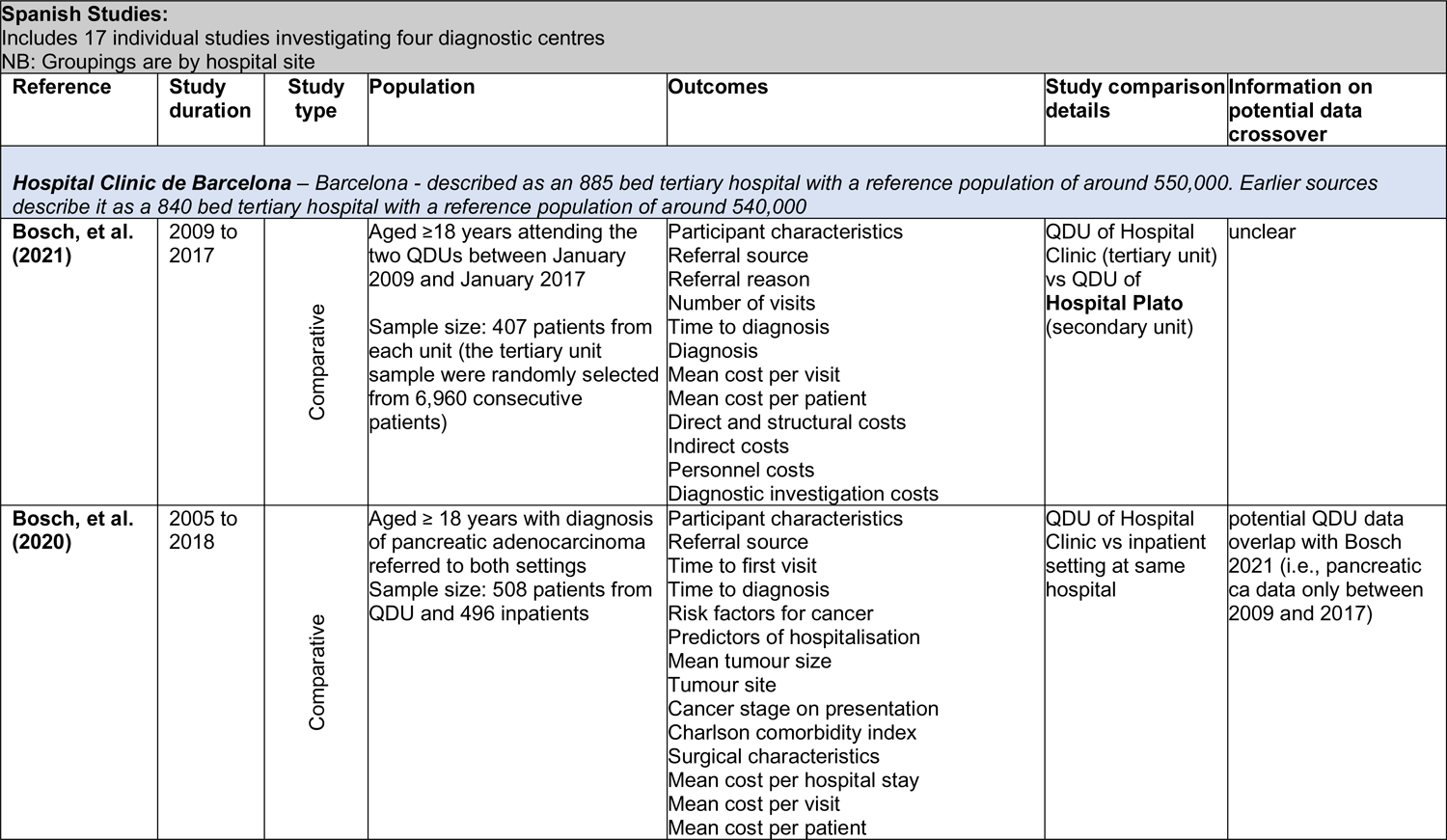
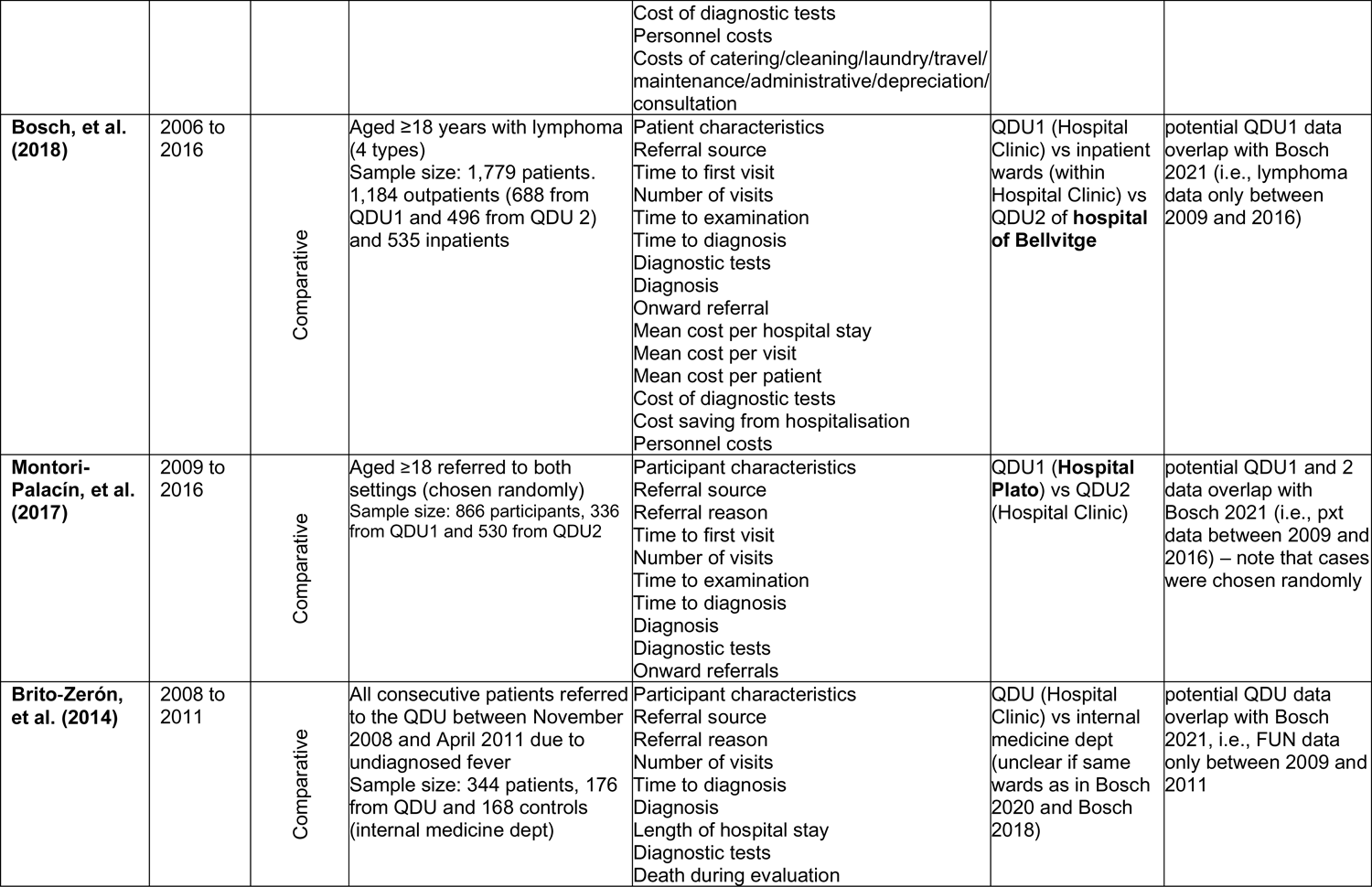
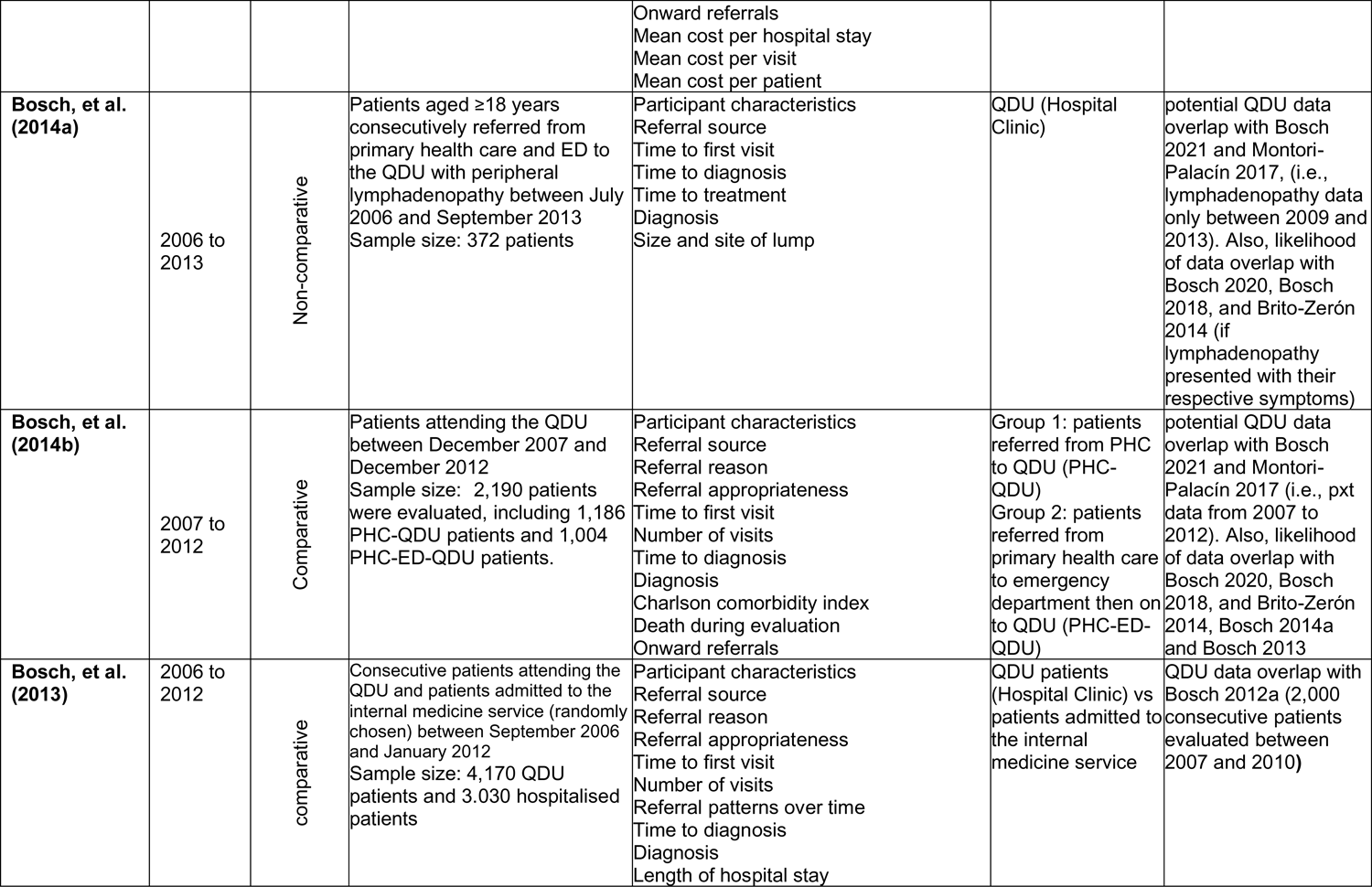
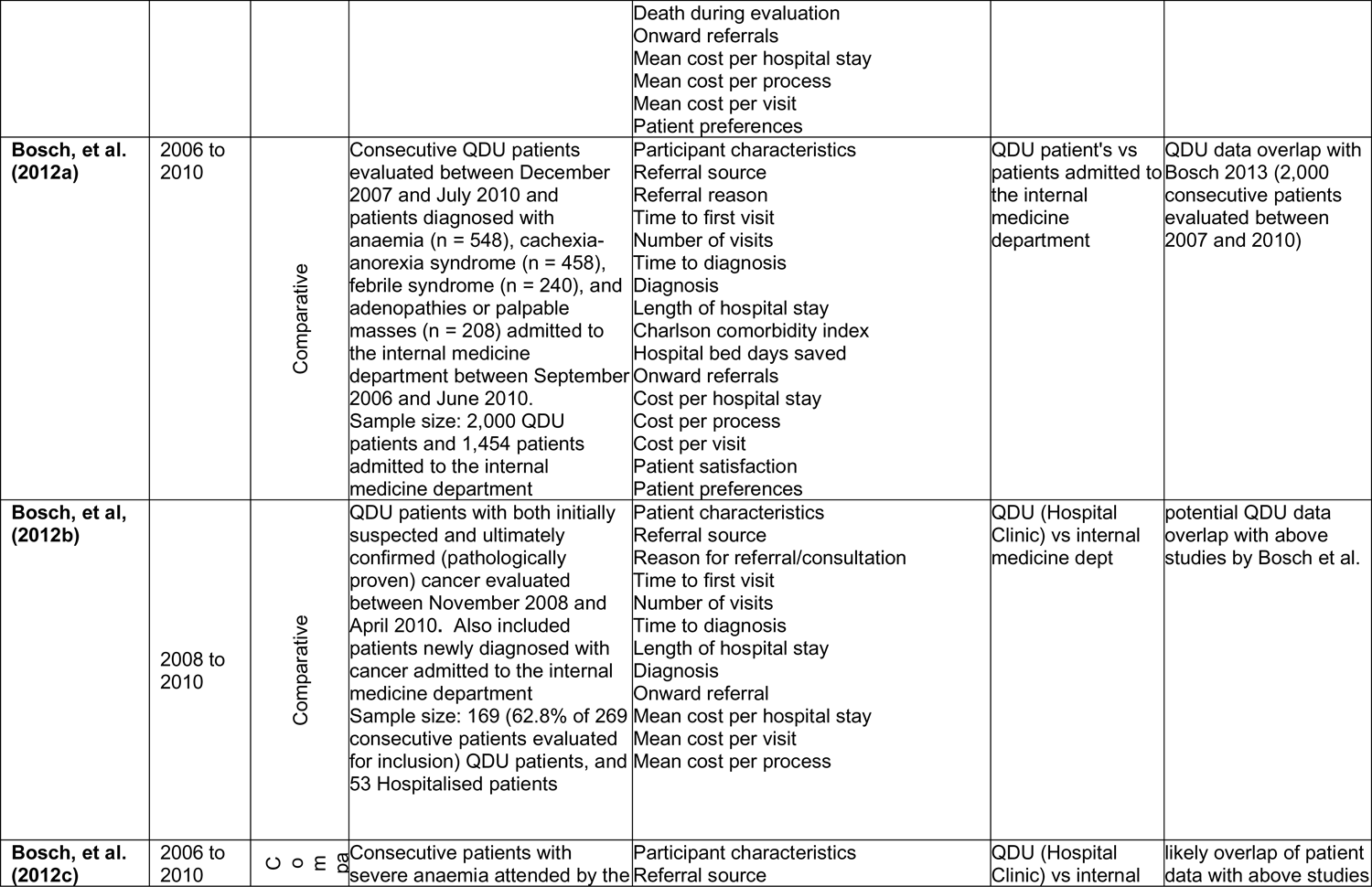
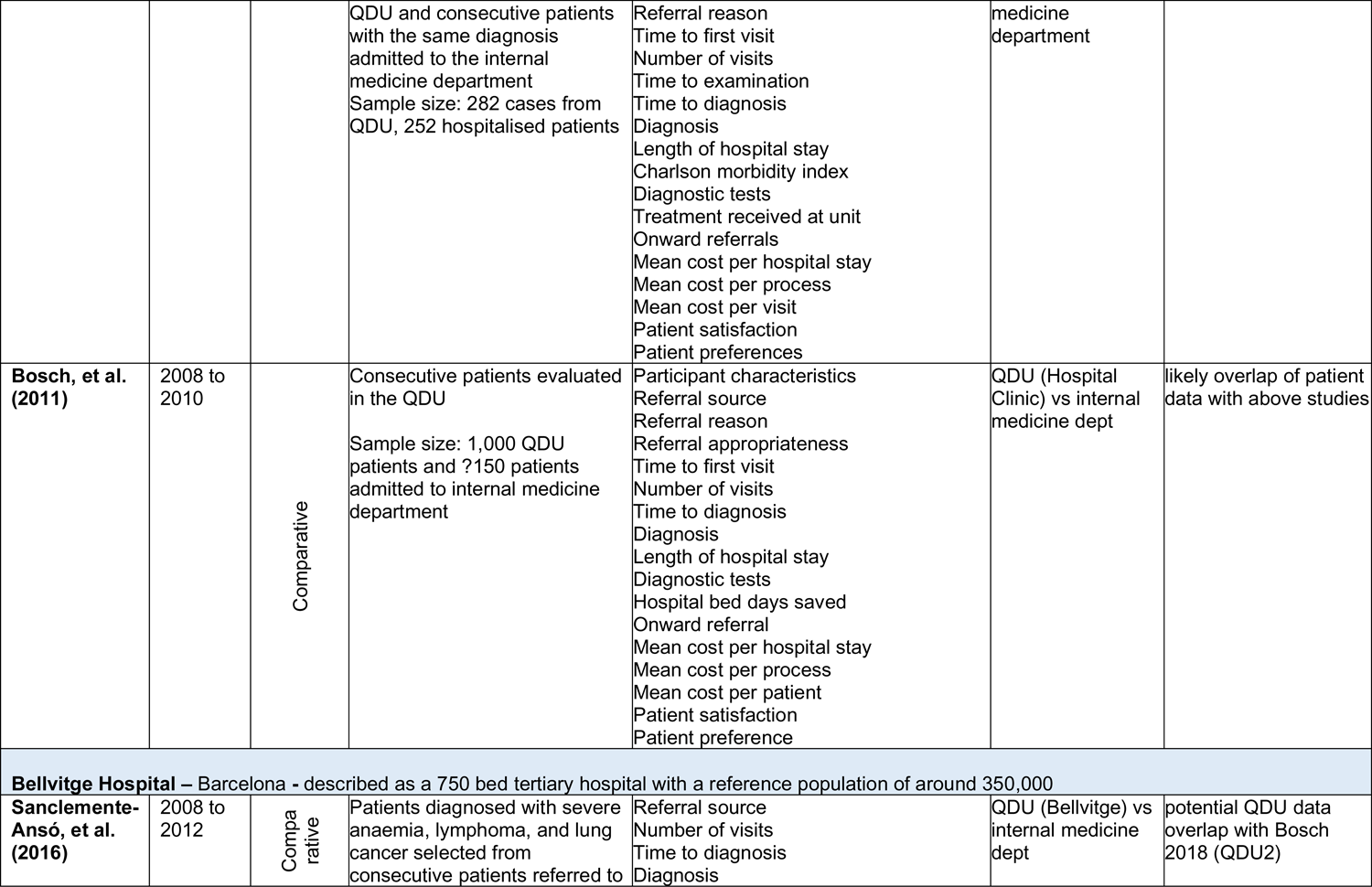
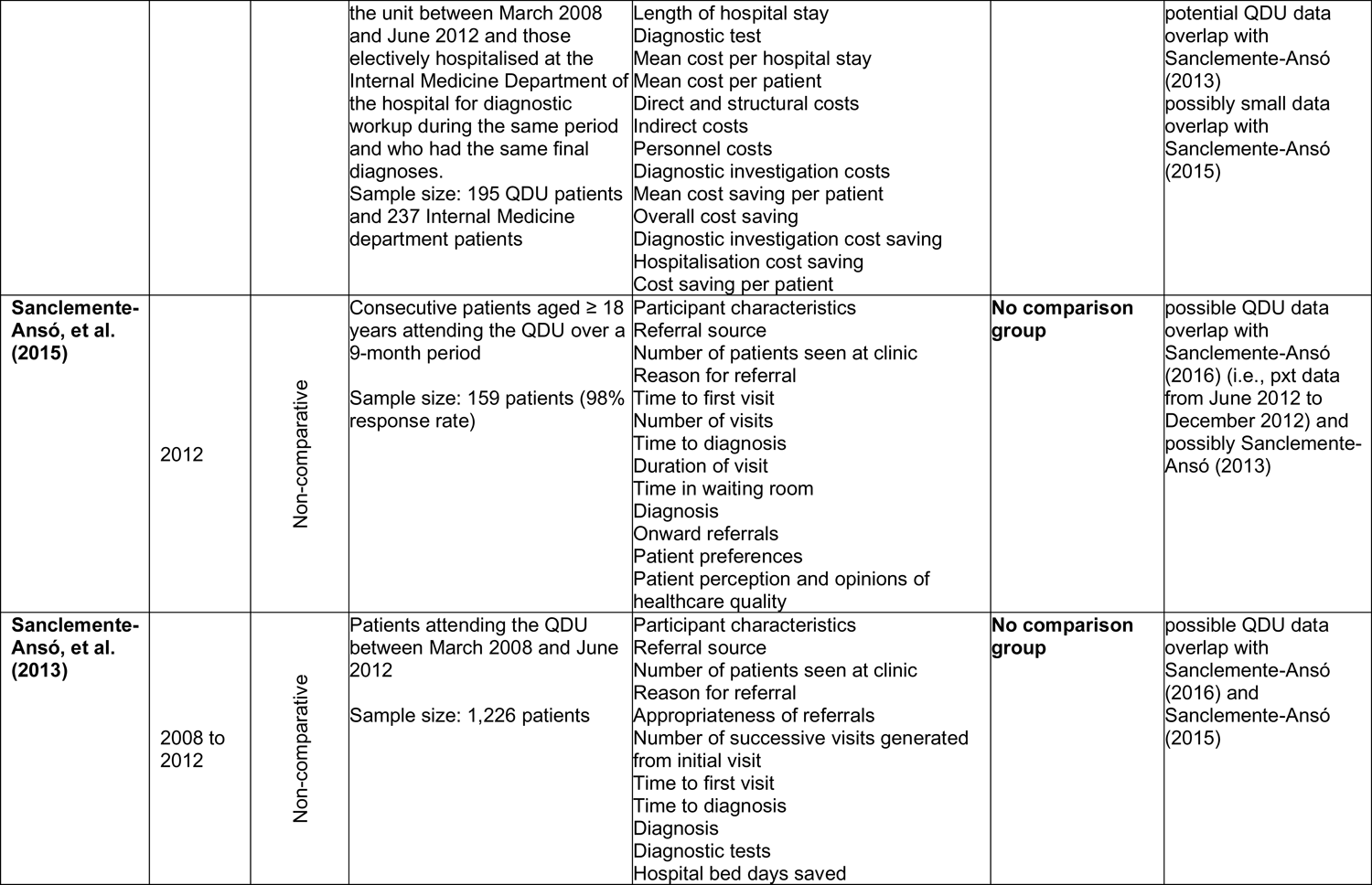
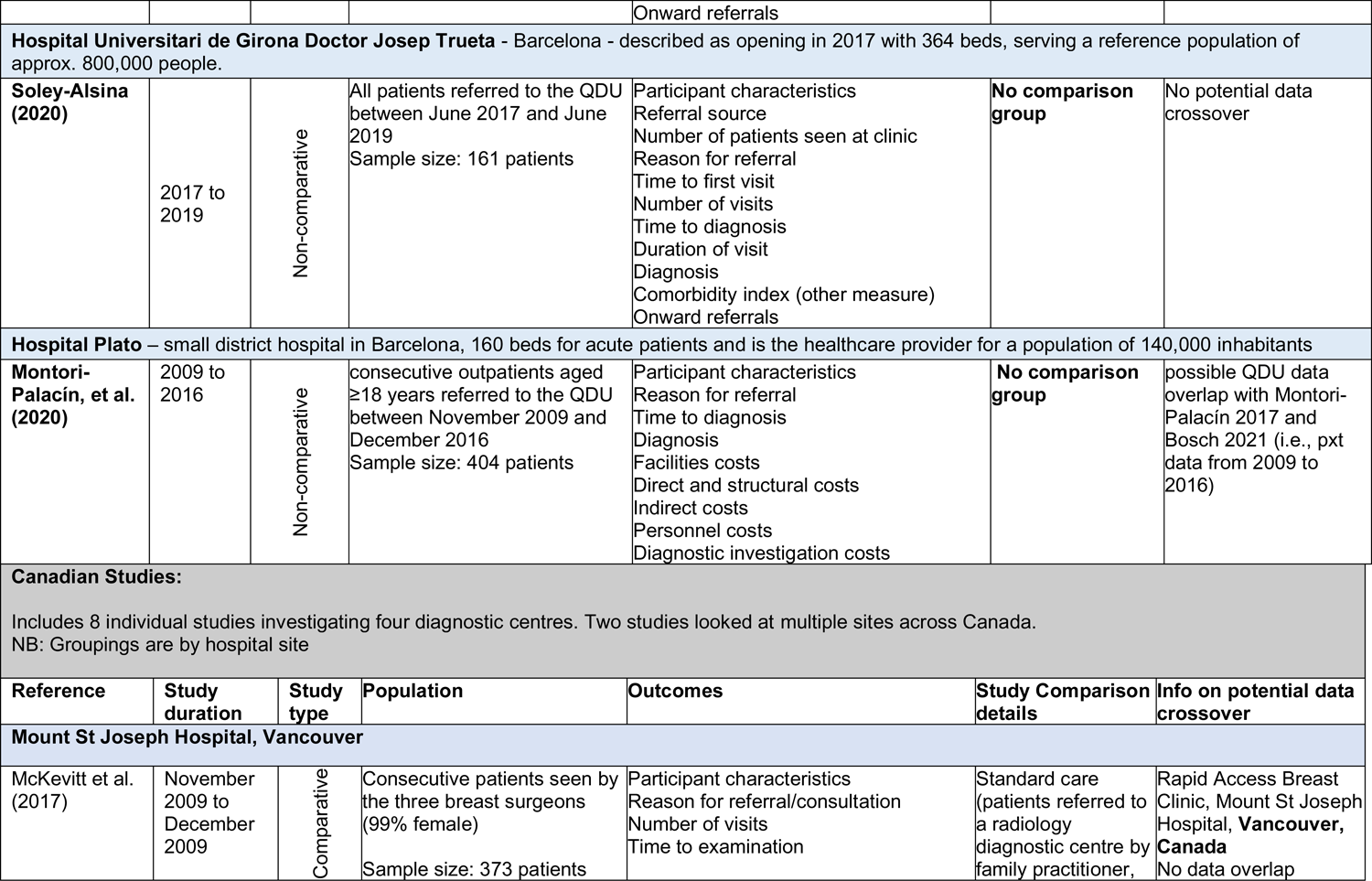
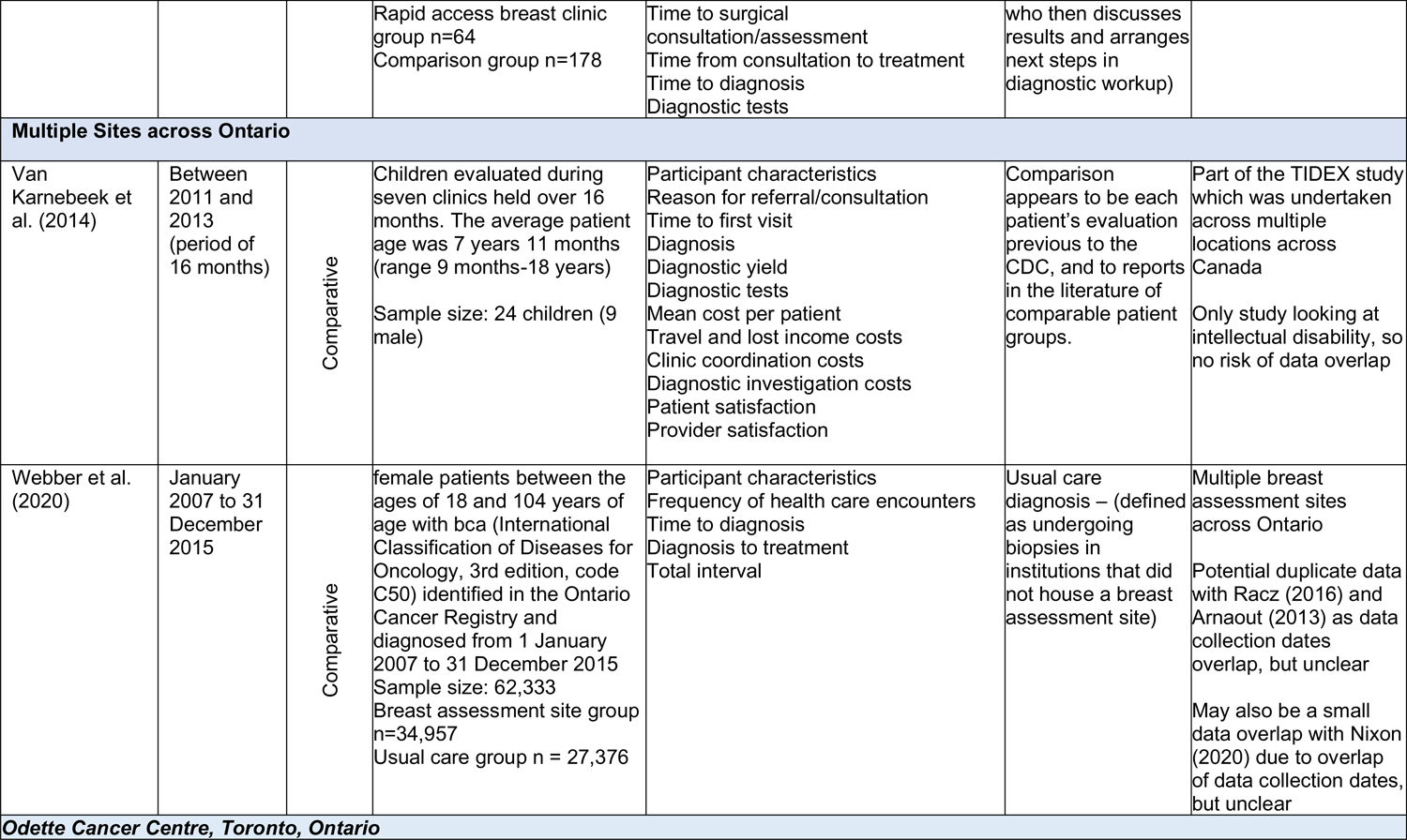
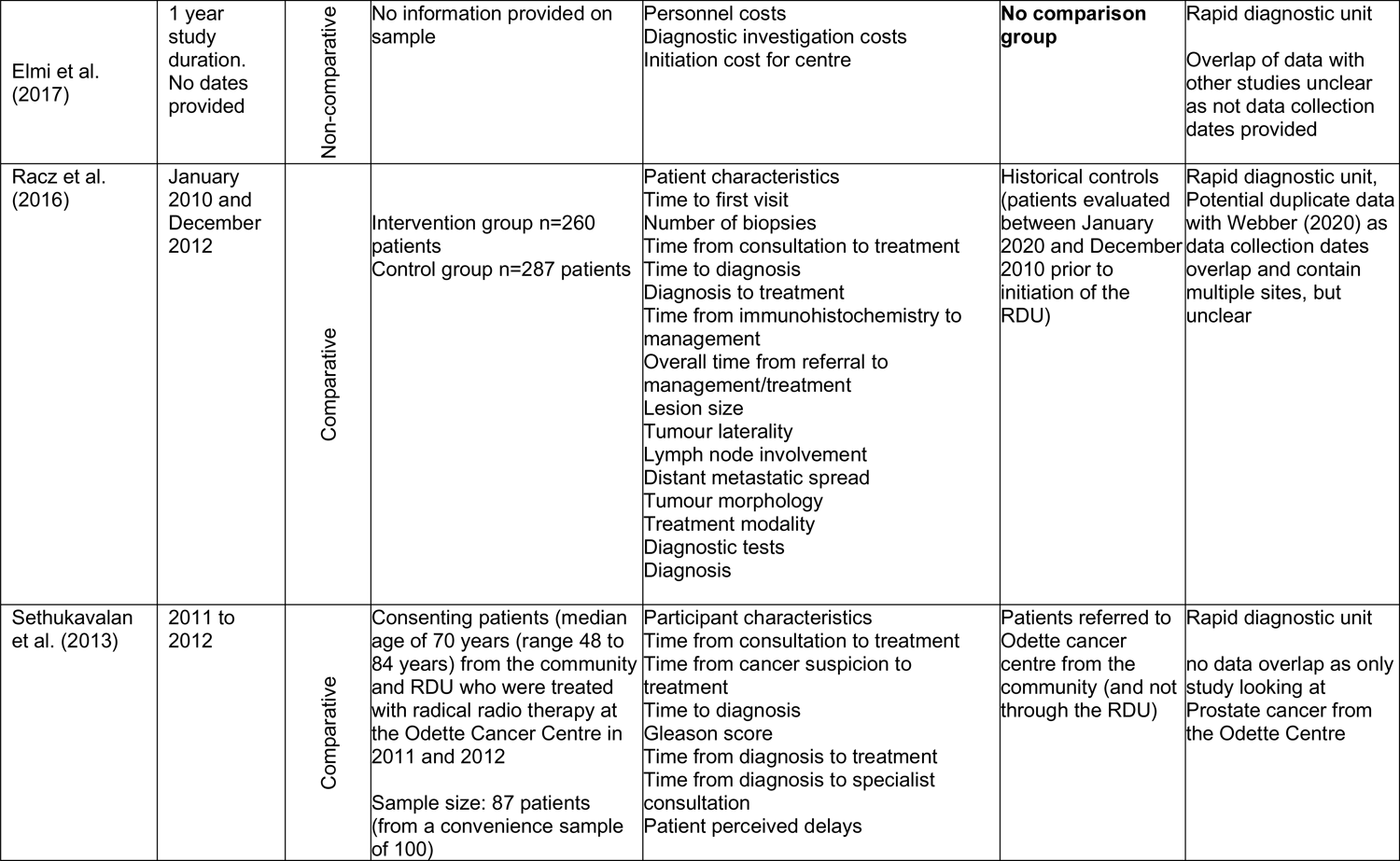
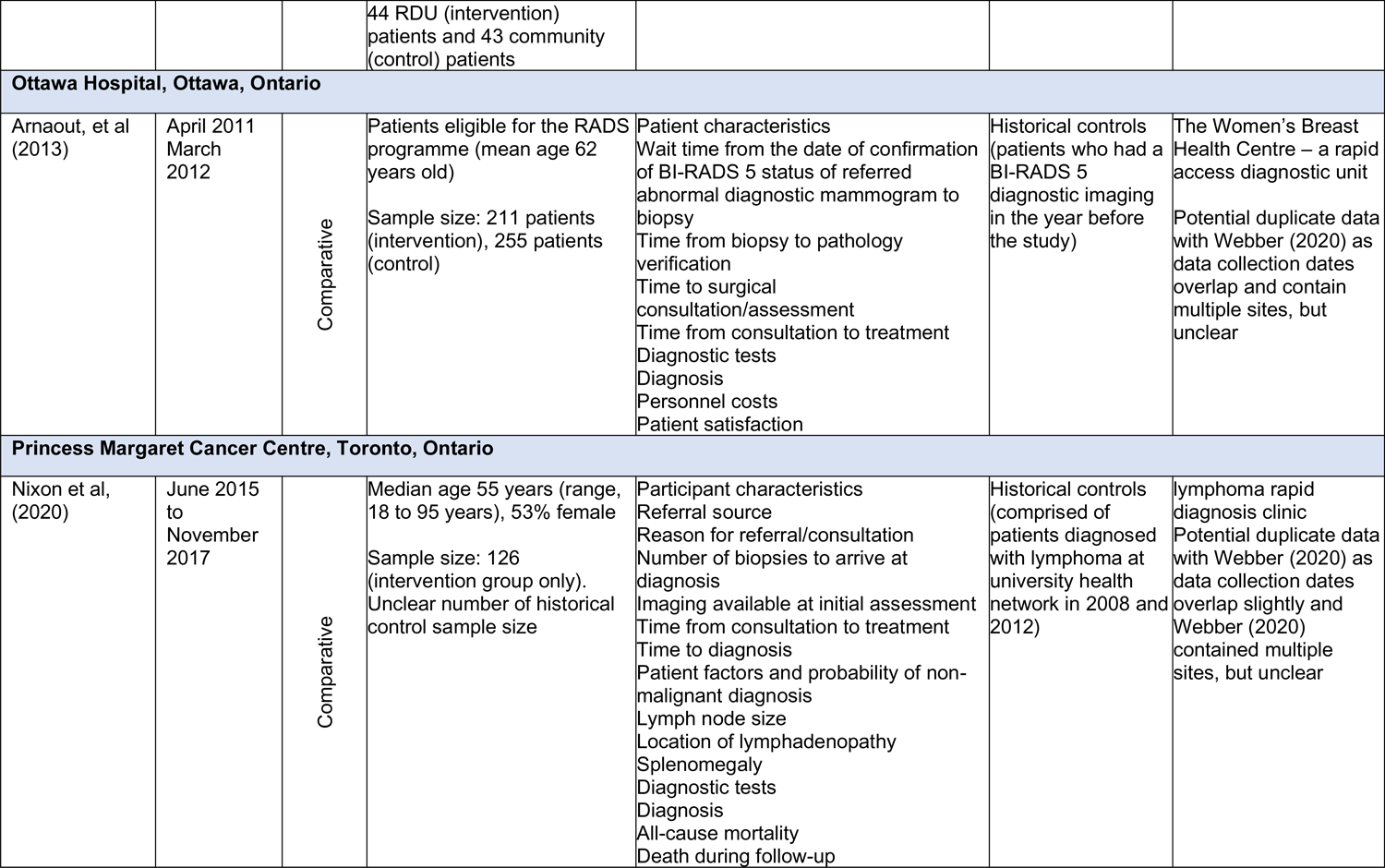
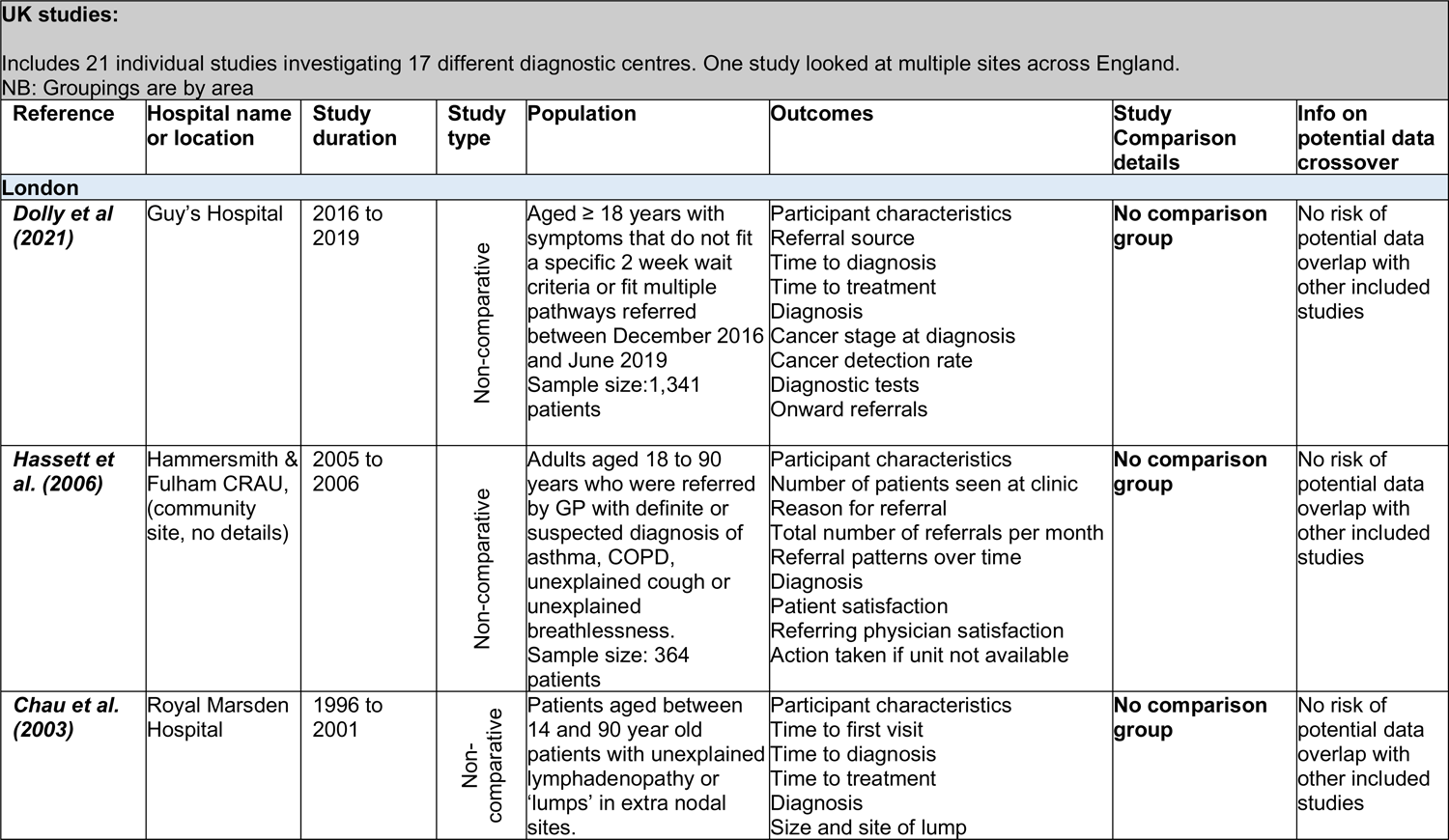
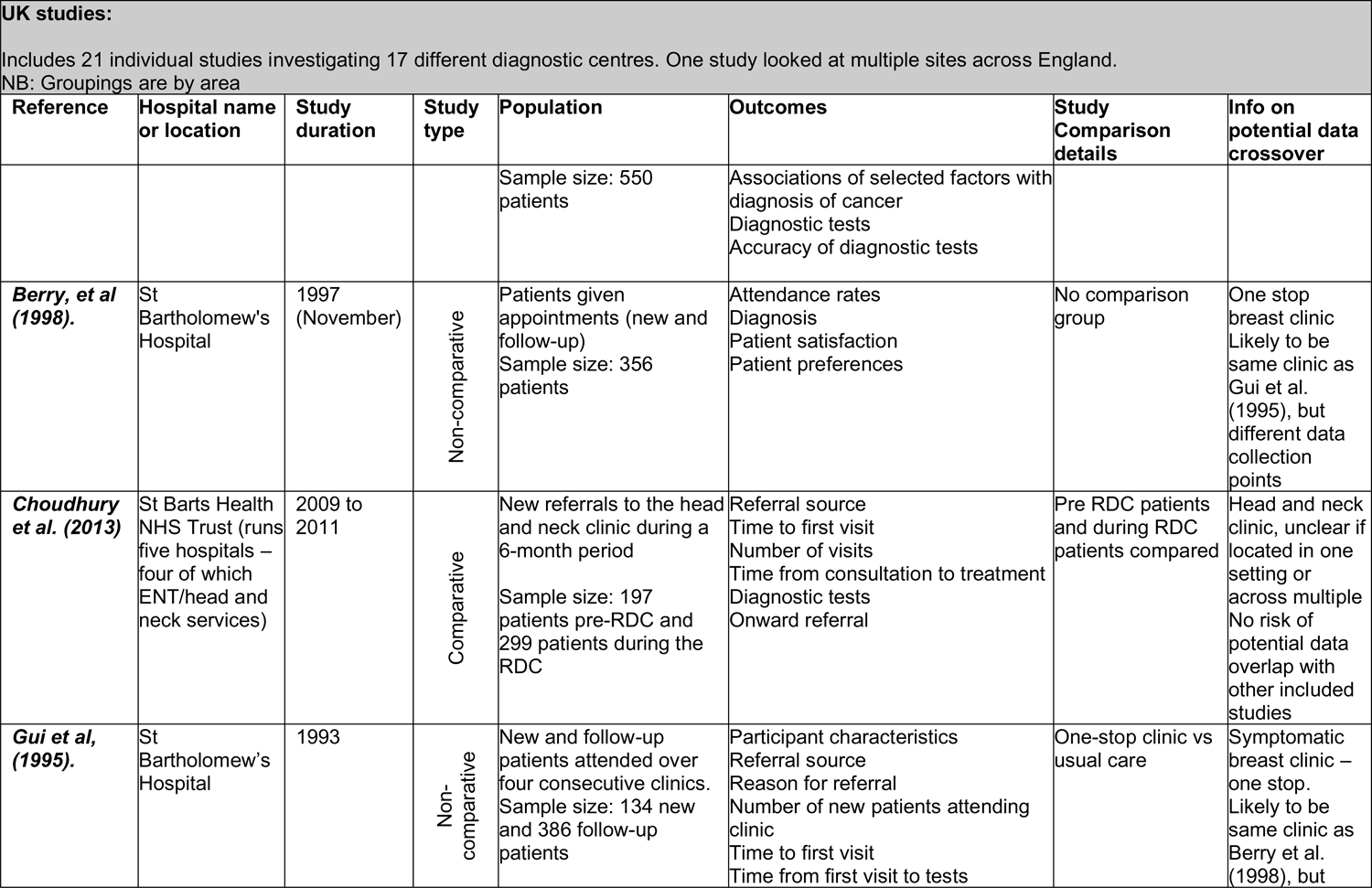
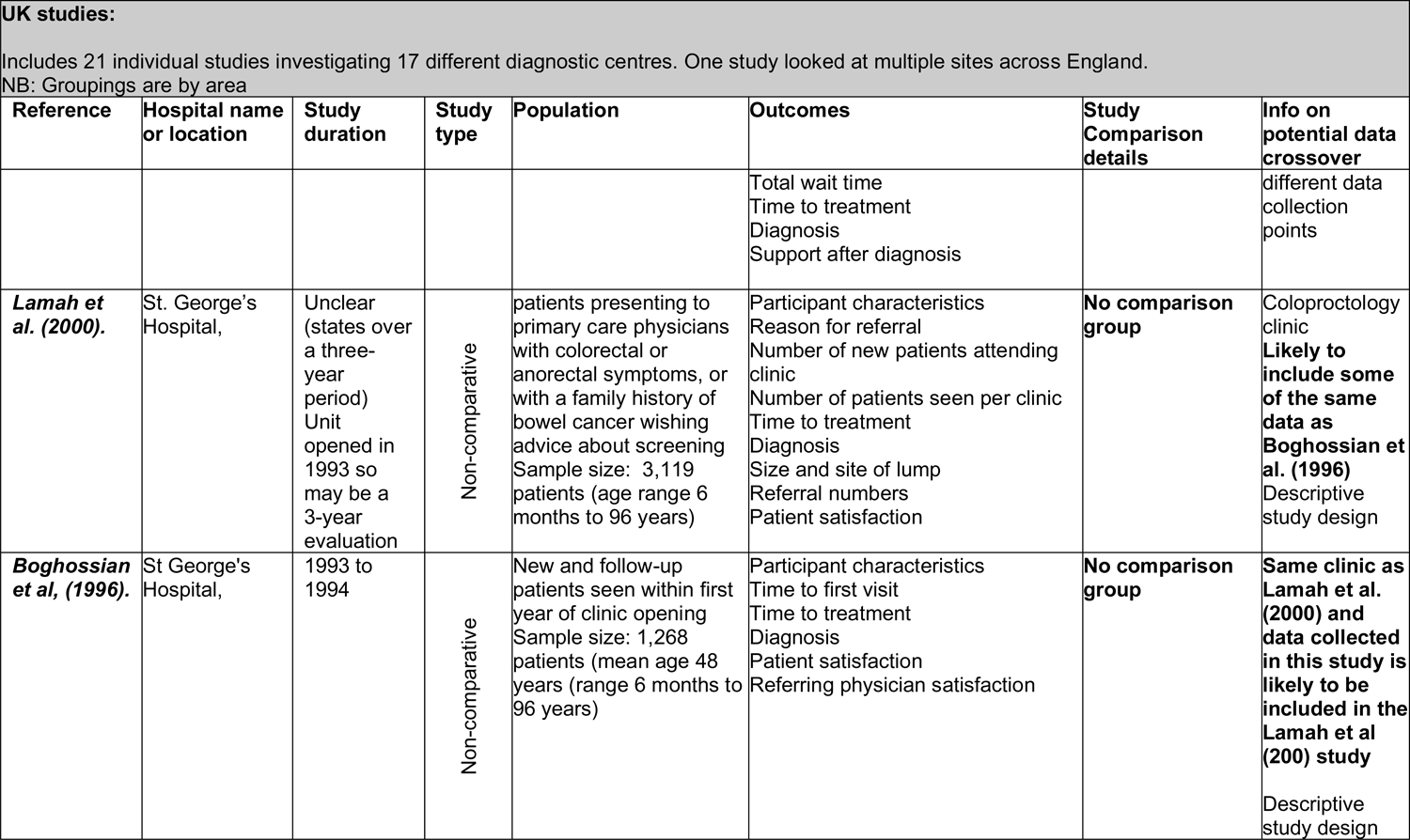
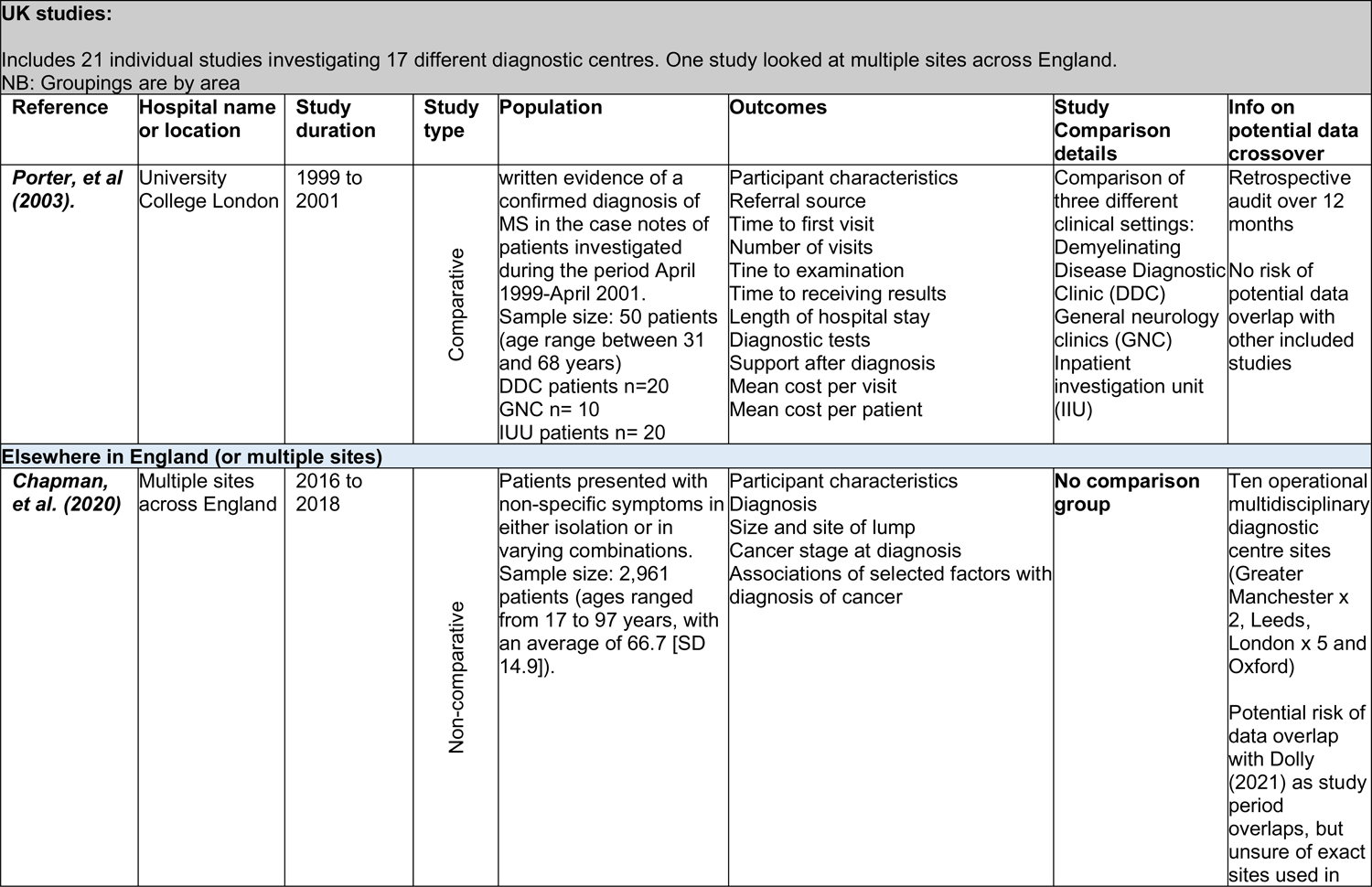
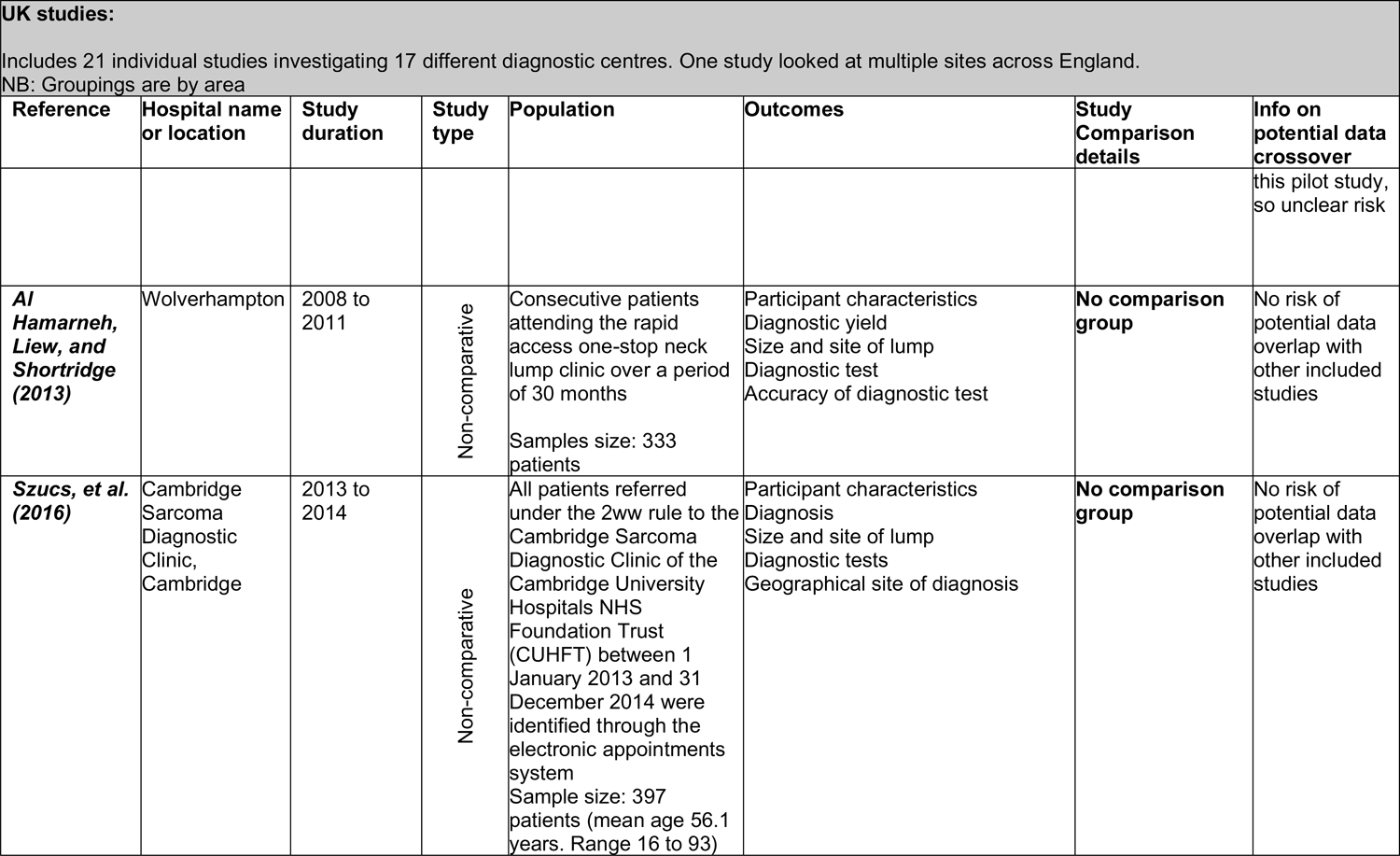
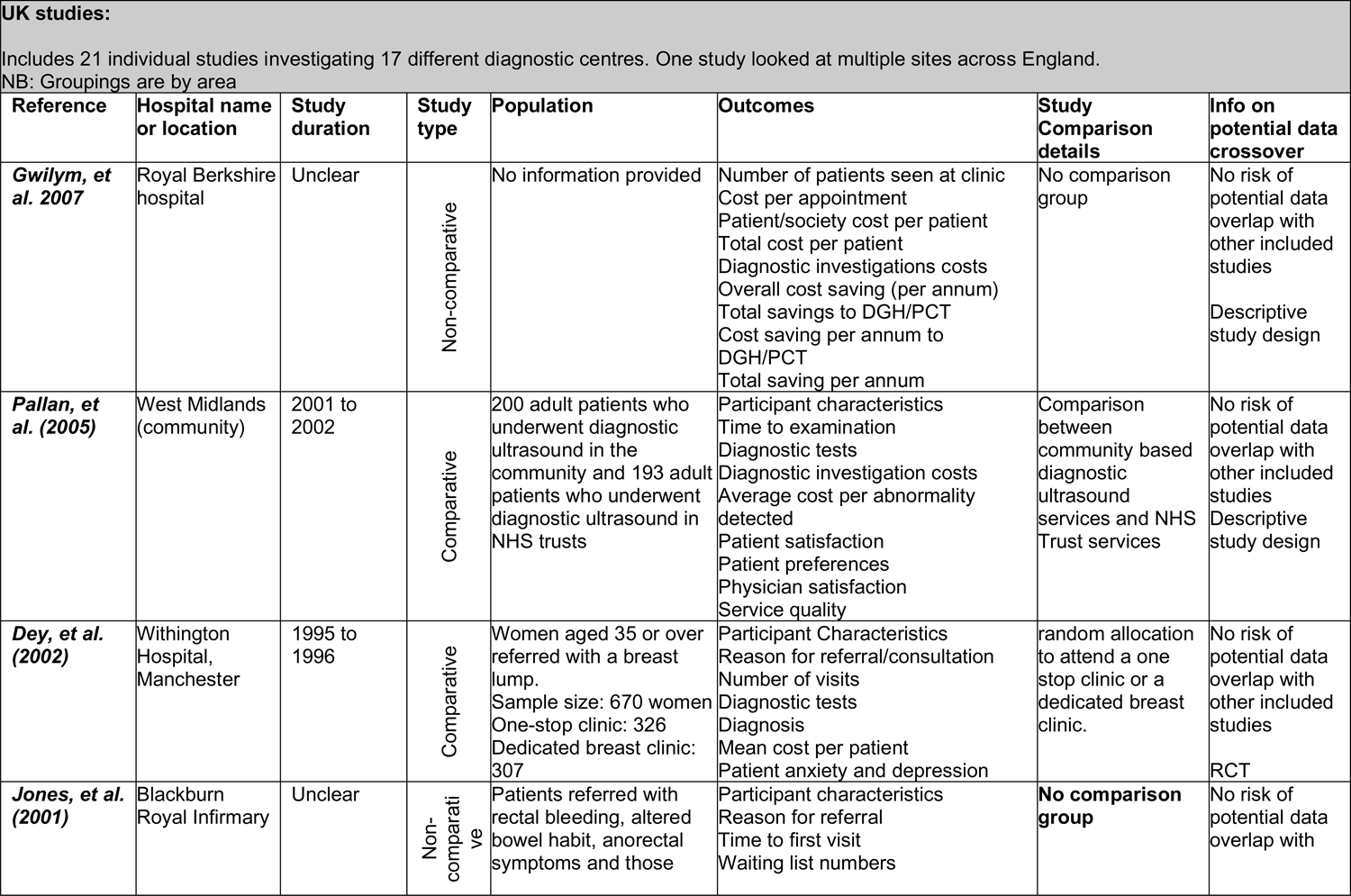
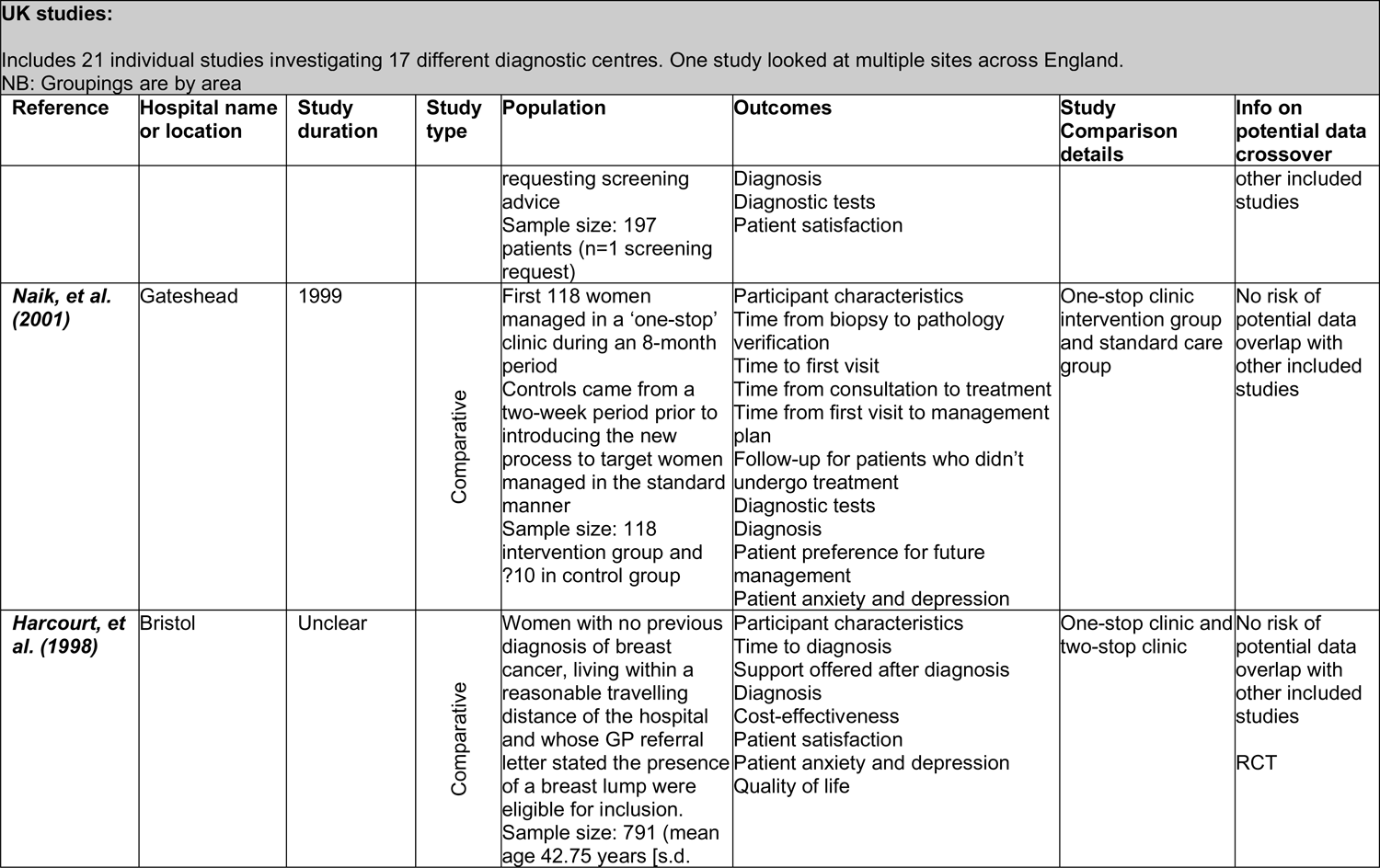
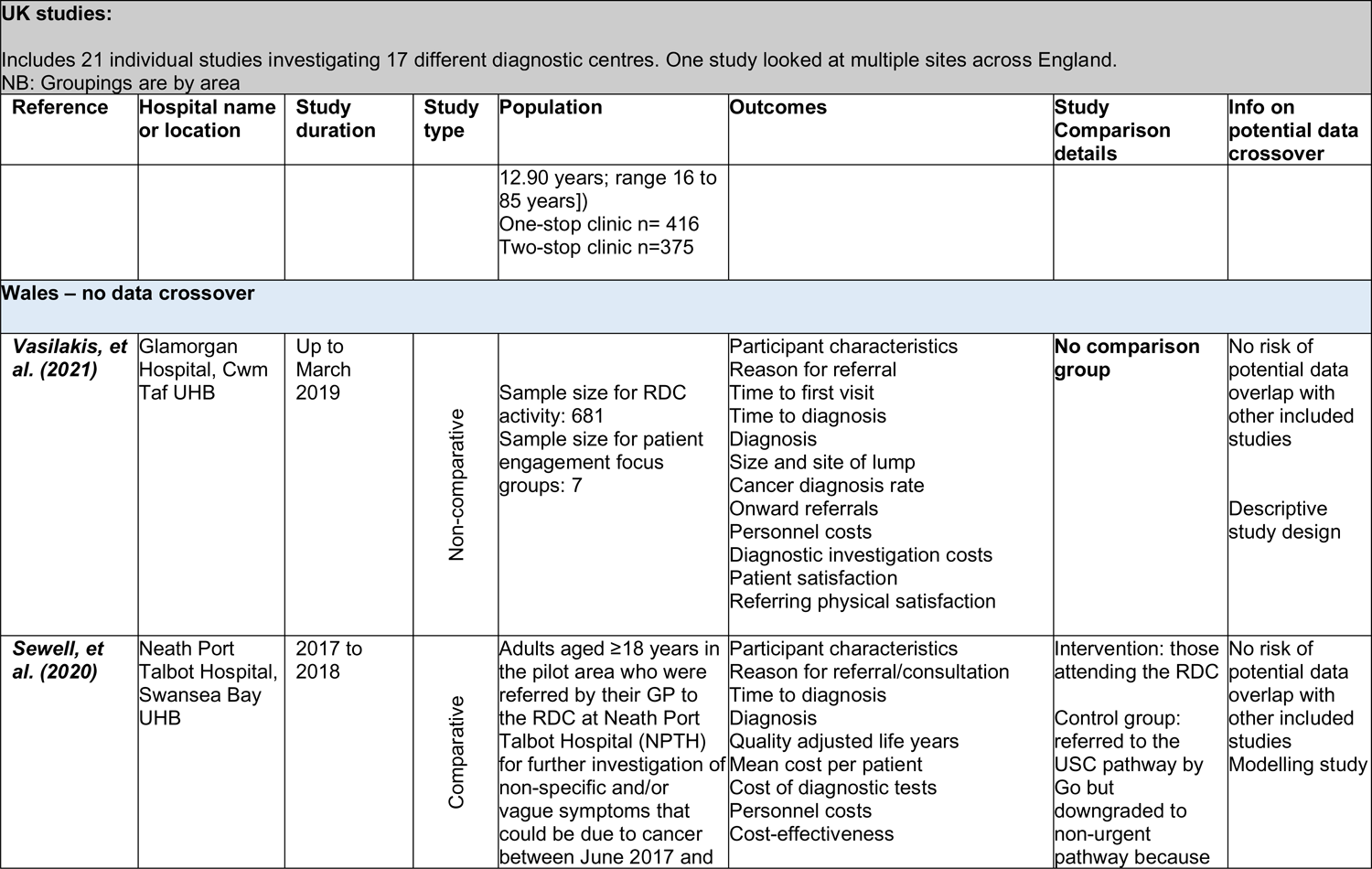
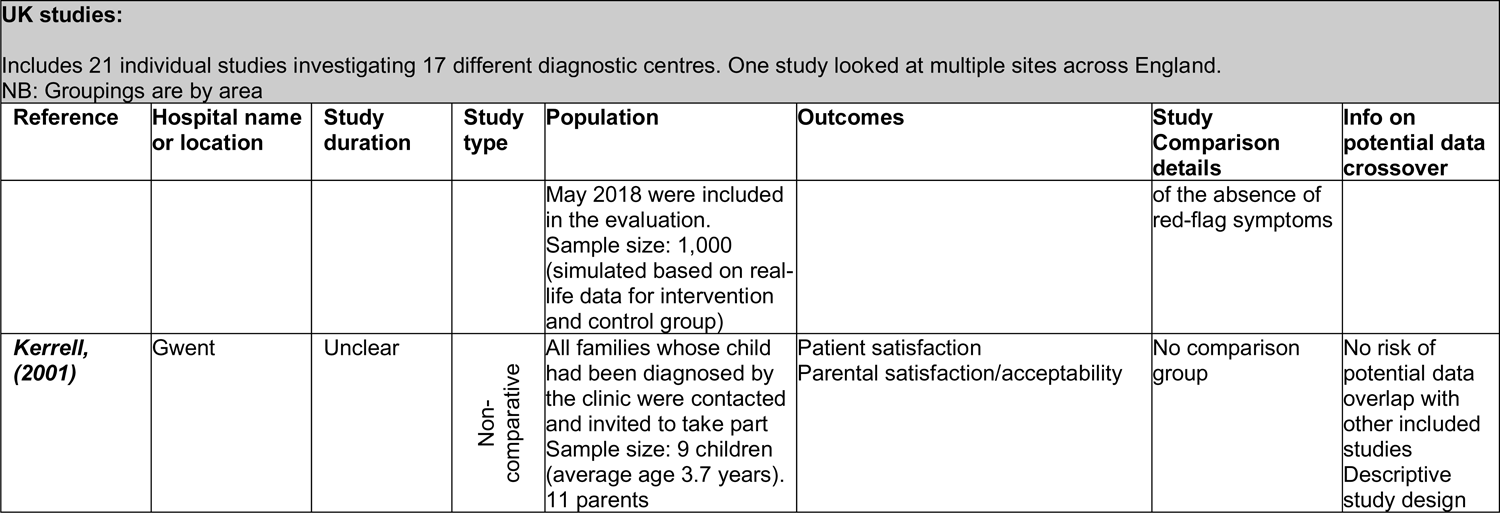
Investigation into the use of the same data across multiple studies

**APPENDIX 3.**
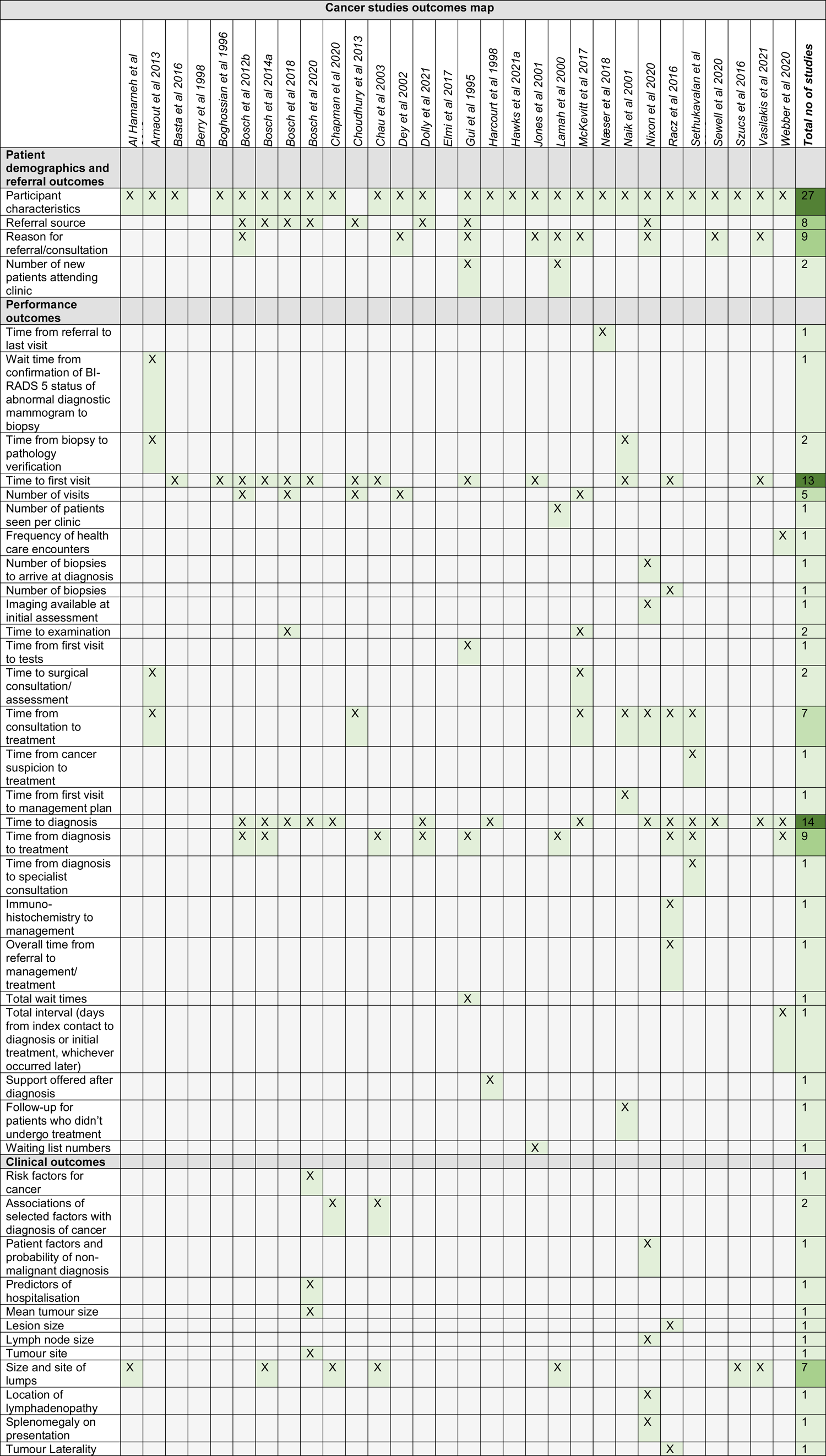
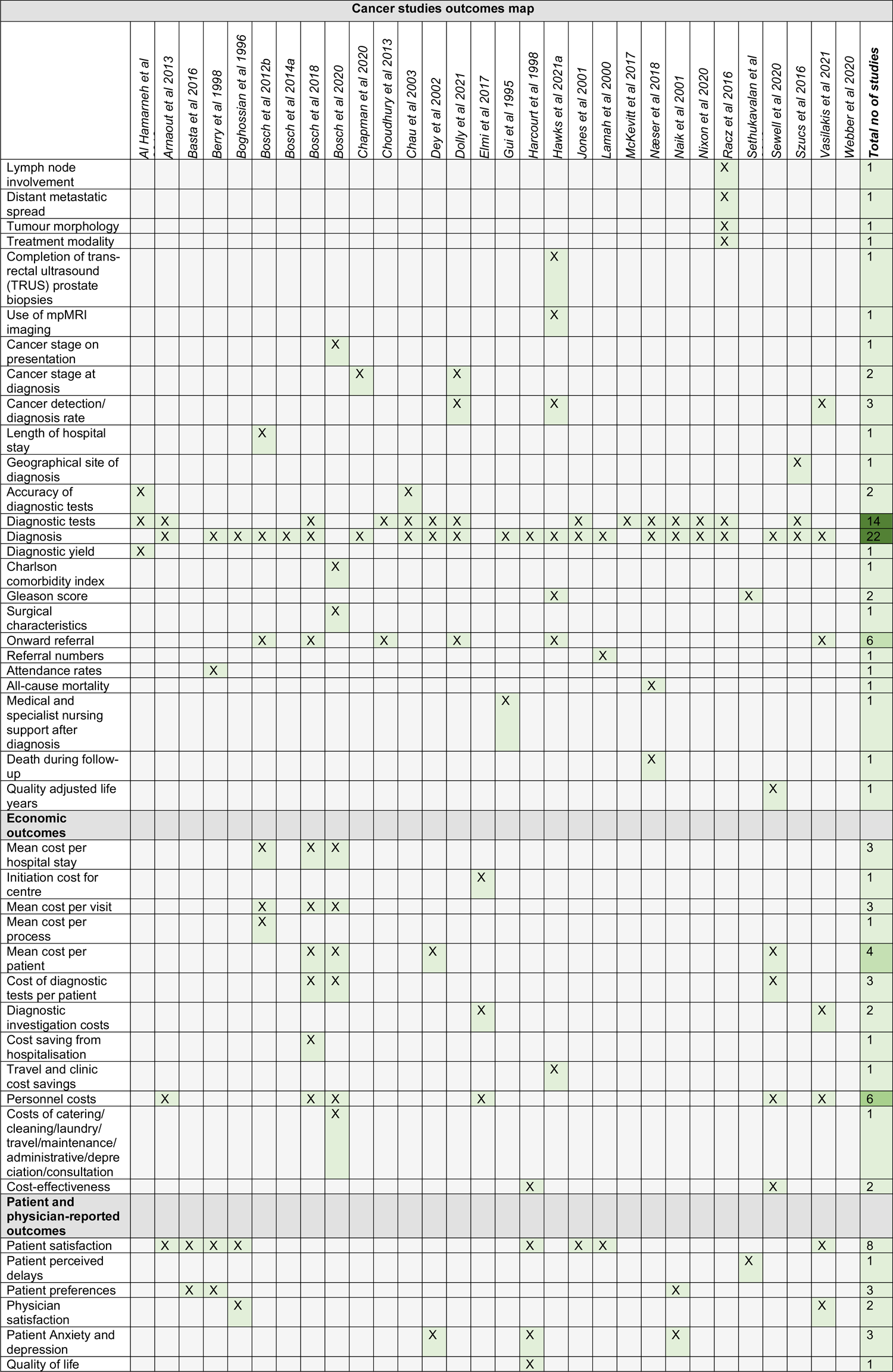
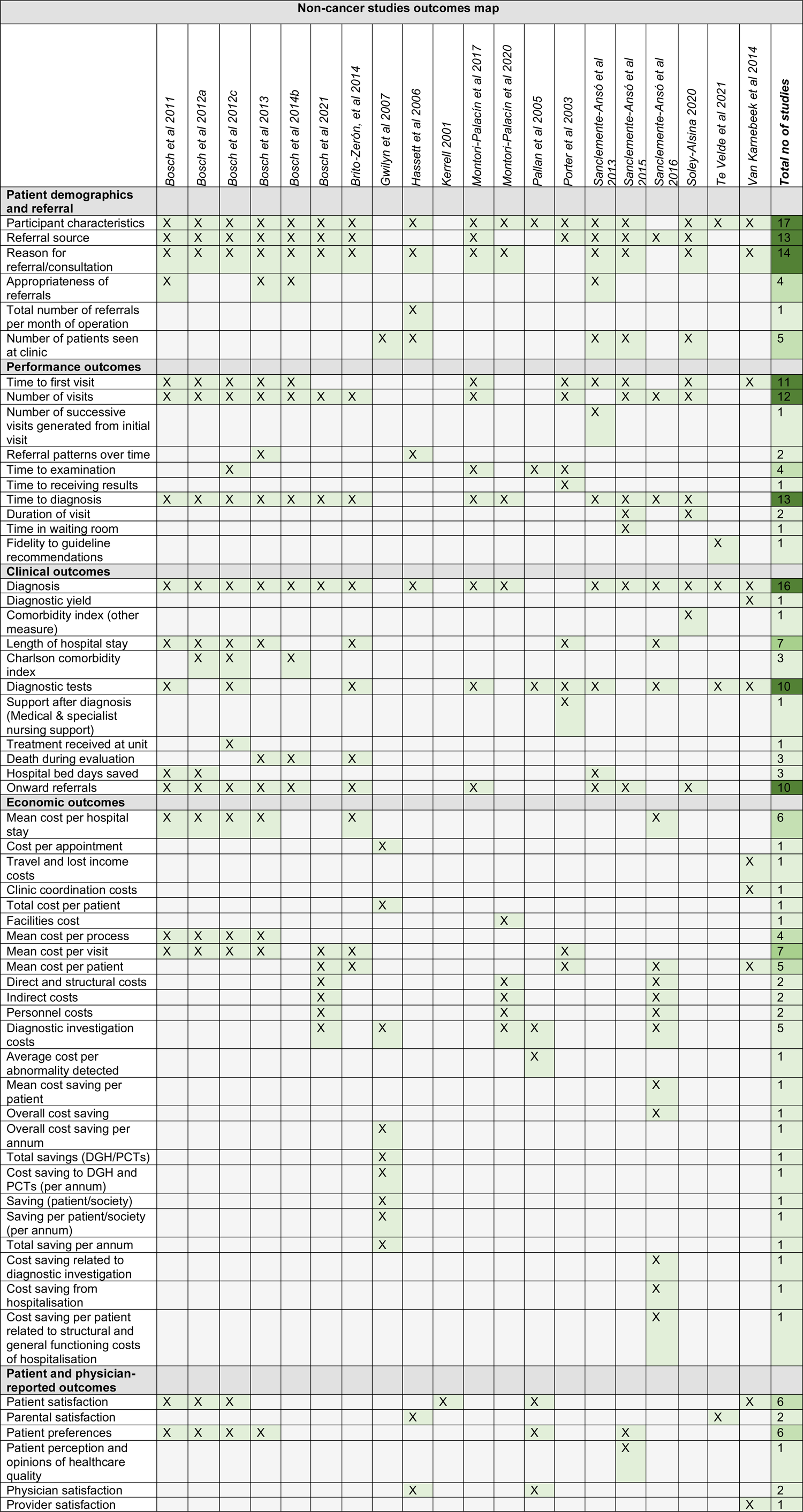
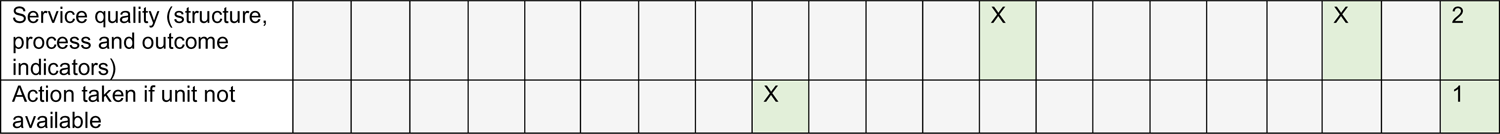
Outcome map for all cancer specific studies and outcome map for all non-cancer specific studies

**APPENDIX 4:**
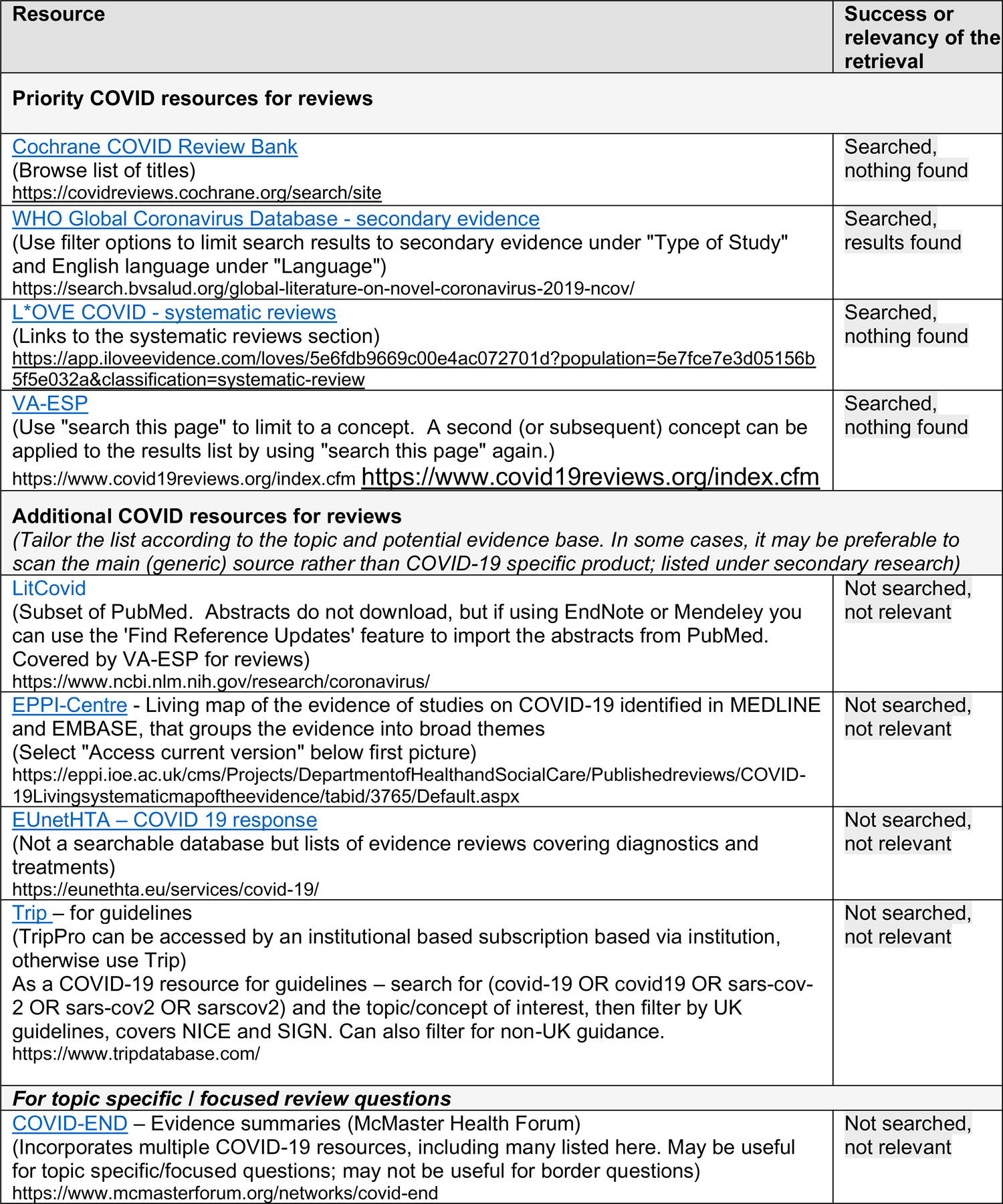
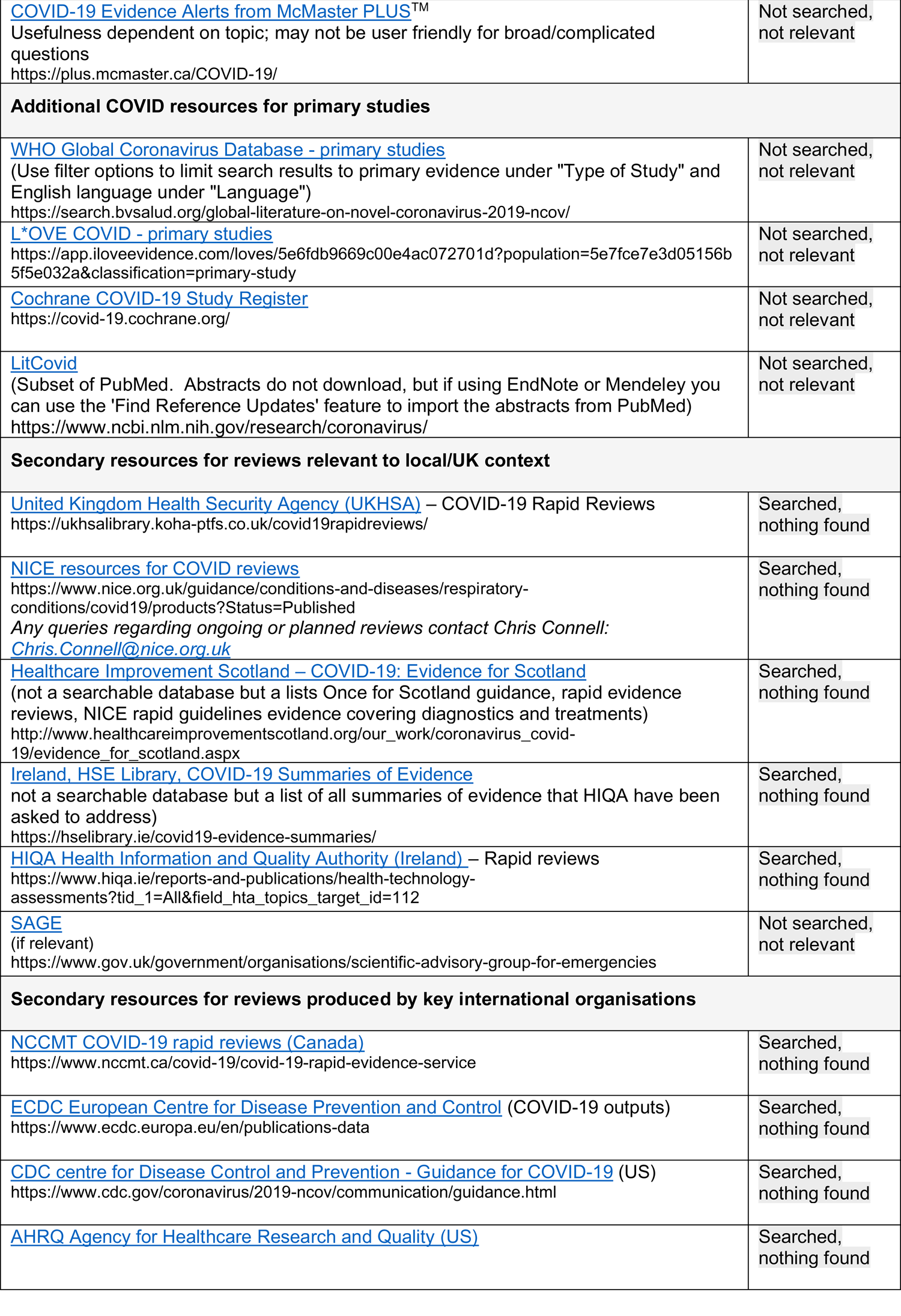
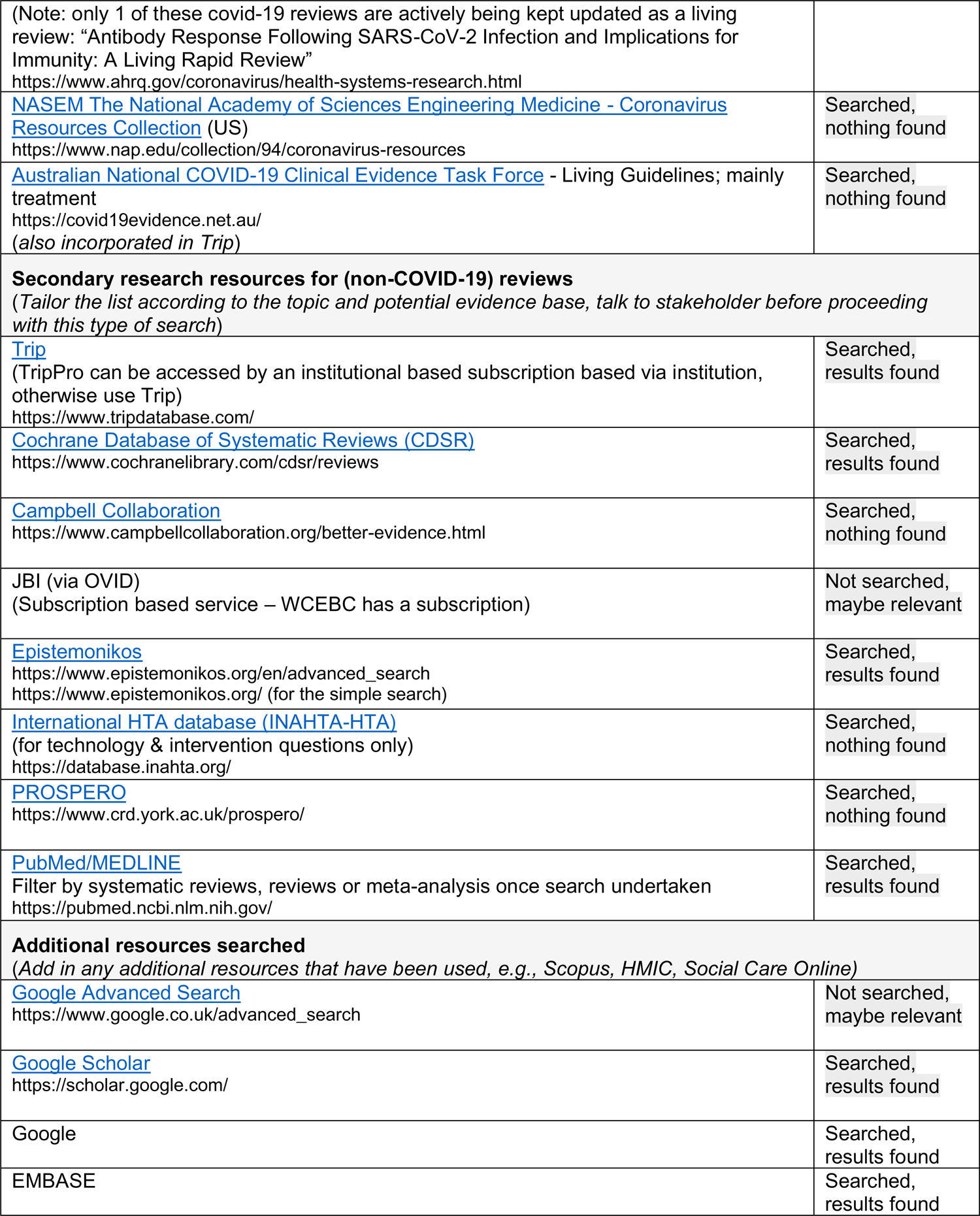
Resources searched during Rapid Review Searching A single list of resources has been developed for guiding and documenting the sources searched as part of a Rapid Review. All ‘core’ resources should be searched, but other resources may be considered if appropriate to the topic, or time allows. For those resources used, record the search strategies used below the table.

**APPENDIX 5.** Search strategy used for MEDLINE

| Set# | Searched for | Results |
| --- | --- | --- |
| S1 | ((TI,AB("community diagnos* centre*"))) | 1 |
| S2 | ((TI, AB ("community diagnos* clinic*"))) | 1 |
| S3 | ((TI, AB ("community diagnos* hub*"))) | 1 |
| S4 | ((TI, AB ("community diagnos* unit*"))) | 0 |
| S5 | ((TI, AB ("rapid diagnos* unit*"))) | 16 |
| S6 | ((TI, AB ("rapid diagnos* clinic*"))) | 12 |
| S7 | ((TI, AB ("rapid diagnos* centre*"))) | 4 |
| S8 | ((TI, AB ("rapid diagnos* hub*"))) | 0 |
| S9 | ((TI, AB ("mobile diagnos* hub*"))) | 0 |
| S10 | ((TI, AB ("mobile diagnos* clinic*"))) | 0 |
| S11 | ((TI, AB ("mobile diagnos* centre*"))) | 0 |
| S12 | ((TI, AB ("mobile diagnos* unit*"))) | 8 |
| S13 | ((TI, AB ("multidisciplinary diagnos* unit*"))) | 0 |
| S14 | ((TI, AB ("multidisciplinary diagnos* hub*"))) | 0 |
| S15 | ((TI, AB ("multidisciplinary diagnos* centre*"))) | 4 |
| S16 | ((TI, AB ("multidisciplinary diagnos* clinic*"))) | 2 |
| S17 | ((TI, AB ("mobile healthcare unit*"))) | 2 |
| S18 | ((TI, AB ("accelerate coordinate evaluate"))) | 3 |
| S19 | S18 OR S17 OR S16 OR S15 OR S14 OR S13 OR S12 OR S11 OR S10 OR S9 OR S8 OR S7 OR S6 OR S5 OR S4 OR S3 OR S2 OR S1 | 52 |
| S20 | ((TI, AB ("diagnos* centre*"))) | 408 |
| S21 | ((TI, AB ("diagnos* clinic"))) | 282 |
| S22 | ((TI, AB ("diagnos* clinics"))) | 86 |
| S23 | ((TI, AB ("diagnos* hub*"))) | 7 |
| S24 | ((TI, AB ("diagnos* unit*"))) | 462 |
| S25 | S24 OR S23 OR S22 OR S21 OR S20 OR S19 | 1247° |
| S26 | (MJMESH.EXACT("Diagnostic Services")) | 1211° |
| S27 | S26 OR S25 | 2436° |

## REFERENCES

1. Chambers, et al (2016). Evidence for models of diagnostic service provision in the community: literature mapping exercise and focused rapid reviews Health Services and Delivery Research, 4(35), pp.1–362.

2. Department of Health and Social Care (2021) Available at: 40 community diagnostic centres launching across England - GOV.UK (www.gov.uk). [Accessed 13 July 2022].

3. Leatherdale; Scott T. Leatherdale (2019) Natural experiment methodology for research: a review of how different methods can support real-world research, International Journal of Social Research Methodology, 22:1, 19–35, DOI: 10.1080/13645579.2018.1488449

4. NHS England (2020) Diagnostic: Recovery and Renewal. Report of the Independent Review of Diagnostic Services for NHS England. Available at: https://www.england.nhs.uk/wp-content/uploads/2020/11/diagnostics-recovery-and-renewal-independent-review-of-diagnostic-services-for-nhs-england-2.pdf [Accessed 12 July 2022].

5. NHS England (2022) One million checks delivered by NHS ‘one stop shops’. News. Available at: NHS England » One million checks delivered by NHS ‘one stop shops’ [Accessed 12 July 2022].

6. NHS (2022) Document 3 - Community Diagnostic Hub (CDH) Draft Qualification Specification. Available at: https://www.ardengemcsu.nhs.uk/media/2585/document-3-cdh-framework-specification-v111.pdf [Accessed 12 July 2022].

7. Welsh Government (2021) NHS activity and performance summary: July and August 2021. Statistics. Available at: https://gov.wales/nhs-activity-and-performance-summary-july-and-august-2021-html [Accessed 12 July 2022].

8. Welsh Government (2022) Our programme for transforming and modernising planned care and reducing waiting lists in Wales. Available at: https://gov.wales/sites/default/files/publications/2022-04/our-programme-for-transforming--and-modernising-planned-care-and-reducing-waiting-lists-in-wales.pdf [Accessed 13 July 2022].

9. Welsh Parliament (2022) Access delayed: The waiting times backlog in NHS Wales. Available at: https://research.senedd.wales/research-articles/access-delayed-the-waiting-times-backlog-in-nhs-wales/ [Accessed 12 July 2022].

